# Genetically modified cones rescue visual function in children

**DOI:** 10.64898/2026.10.13.26363794

**Authors:** Roni O Maimon-Mor, Mahtab Farahbakhsh, Andrew T. Rider, Andrew Stockman, Téo Berthy-Parker, Mohamed M Katta, Michel Michaelides, Tessa M Dekker

## Abstract

Gene therapy can restore retinal signals absent since birth in animal models, yet the translation to humans remains challenging. An unresolved possibility is that meaningful retinal rescue already occurs in humans, but its impact is masked by downstream neural bottlenecks shaped by long-term deprivation. Tracing restored signals through the visual system is therefore essential to distinguish rescue and constraints.

We address this challenge in *CNGA3*-associated achromatopsia, in which retinal cone photoreceptors are present but non-functional from birth. We used tests specifically targeting cone-mediated processing to trace signals from genetically modified cones from the retina to visual cortex and perception.

After treatment in late childhood, we find clear evidence of new cone signals reaching visual cortex in 4 out of 7 patients. These signals were replicable over years, retinotopically organised, available for conscious perception and showed higher temporal resolution. Recovery of foveal acuity and canonical colour vision remained limited, consistent with neuroplasticity constraints on specialised neurocircuitry beyond critical developmental windows.

Together, these findings show that the human brain can incorporate a new sensory signal after years of complete congenital deprivation, while revealing selective bottlenecks that limit full visual recovery. They provide a framework for optimising sensory restoration: measure rescued input, trace downstream integration of signal, and tailor treatment timing and rehabilitation to maximise functional gain.

## Introduction

Gene therapies are revolutionary technologies that aim to eradicate the underlying causes of serious heritable diseases by introducing a functional gene into target tissue. To develop gene therapies for nervous system disorders such as sight loss (Fuller-Carter et al., 2020; Georgiou et al., 2021; Trapani & Auricchio, 2018), repairing local tissue is not enough. Recovery of function relies not only on the recovery of retinal signalling, but also on complex brain-wide networks. A central challenge for sight restoration is whether neural circuits shaped by lifelong atypical input can integrate novel signals from a genetically modified retina.

Here, we address this question in people with achromatopsia, a congenital retinal disorder in which cone photoreceptors are non-functional from birth (Aboshiha et al., 2014; Hirji, Aboshiha, et al., 2018). Cones enable high-detail and colour vision in daylight, whereas rods support low-light and peripheral vision. People with complete achromatopsia rely exclusively on rod-mediated input, resulting in low spatial and temporal acuity (“blurry” and “slower” vision), complete colour blindness, extreme light sensitivity (“day blindness”), pendular nystagmus, and a central blind-spot due to the rod-free fovea. Most achromatopsia cases are caused by disease-causing variants in *CNGA3* or *CNGB3* genes that encode for cone-specific components of the phototransduction cascade. As cones are present but non-functional (Georgiou et al., 2019; Hirji, Georgiou, et al., 2018), it makes achromatopsia a strong model for sight rescue as gene therapy can, in principle, re-establish signalling in the existing cones. At the same time, because cone-driven input has been absent since birth, the condition provides a stringent test of whether newly reinstated cone signals can propagate through the visual system and be integrated by downstream pathways to support previously unavailable percepts.

In animal models of achromatopsia, gene therapy has restored cone function and supported photopic visual behaviour (e.g., navigation), with some mouse studies also showing improved visual acuity and light sensitivity (Alexander et al., 2007; Banin et al., 2015; Carvalho et al., 2011; Gootwine et al., 2017; Hassall et al., 2017; Komáromy et al., 2010; Michalakis et al., 2010; Pang et al., 2012; Zhang et al., 2019). Yet, despite these successes, there is currently no approved human treatment. Several clinical trials have produced modest and inconsistent gains on standard endpoints such as acuity, contrast sensitivity, photoaversion and colour vision, leading to stalled progress in this field (Fischer et al., 2020; Michaelides et al., 2023; Reichel et al., 2022). We hypothesised that this translational gap does not necessarily reflect a failure of gene therapy to rescue cone signalling at the retina. Rather because standard behavioural endpoints may conflate two separate processes: the restoration of cone-driven retinal input and the ability of downstream visual circuitry to use that input effectively, making it difficult to identify which stage limits recovery. Consequently, a gene therapy may successfully introduce cone-driven input at the retina, while functional gains in standard measures remain limited by neuroplasticity bottlenecks in a visual system developed with only rod input.

To test this we followed seven individuals with *CNGA3*-associated achromatopsia who received monocular subretinal AAV gene therapy in their worse eye (NCT02935517), between ages 8 and 15 years (Table 1). In an observational study alongside the trial, we used converging fMRI and psychophysical measures designed to isolate cone-mediated processing, to trace functional rescue across processing stages. We asked two questions: 1. Do signals from genetically modified cones emerge and reach visual cortex? 2. Which specialised neural pathways can use these previously unavailable signals to support visual functions?

**Table 1.**
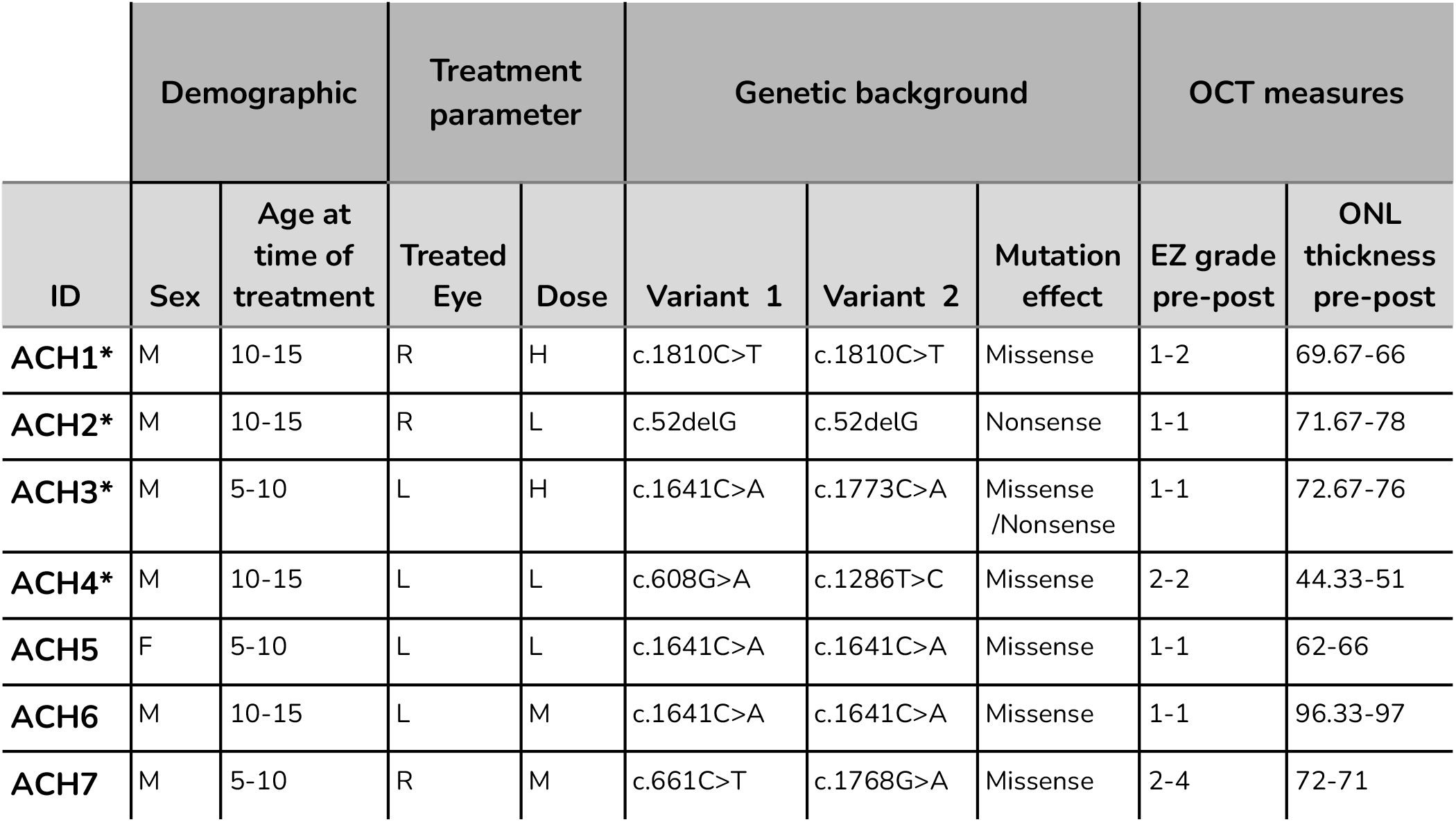
Patients’ background, treatment and retinal details. Sex: M-male, F-female. Eye: R-right, L-left. Dose: H-high, M-intermediate, L-low. OCT: post measures are for the last available post for each patient. Patients showing a clear treatment effect are marked with*

|  | Demographic |  | Treatment parameter |  | Genetic background |  |  | OCT measures |  |
| --- | --- | --- | --- | --- | --- | --- | --- | --- | --- |
| ID | Sex | Age at time of treatment | Treated Eye | Dose | Variant 1 | Variant 2 | Mutation effect | EZ grade pre-post | ONL thickness pre-post |
| ACH1* | M | 10-15 | R | H | c.1810C>T | c.1810C>T | Missense | 1-2 | 69.67-66 |
| ACH2* | M | 10-15 | R | L | c.52delG | c.52delG | Nonsense | 1-1 | 71.67-78 |
| ACH3* | M | 5-10 | L | H | c.1641C>A | c.1773C>A | Missense /Nonsense | 1-1 | 72.67-76 |
| ACH4* | M | 10-15 | L | L | c.608G>A | c.1286T>C | Missense | 2-2 | 44.33-51 |
| ACH5 | F | 5-10 | L | L | c.1641C>A | c.1641C>A | Missense | 1-1 | 62-66 |
| ACH6 | M | 10-15 | L | M | c.1641C>A | c.1641C>A | Missense | 1-1 | 96.33-97 |
| ACH7 | M | 5-10 | R | M | c.661C>T | c.1768G>A | Missense | 2-4 | 72-71 |

In four of seven patients, we find converging evidence that gene therapy has rescued robust cone-mediated signalling in visual cortex, absent before treatment, which extends our previous case study (Farahbakhsh et al., 2022). This rescue persists across years and enables perception of previously invisible stimuli. It induces cone-specific changes in spectral sensitivity and perceived brightness and supports faster visual processing - functional gains that only cones can provide. However, rescue is also selective. We find limited acuity gains and link these to weak rescue of foveal signalling to visual cortex. Although some patients can detect colour cues, colour vision remains atypical with no evidence of rescued cone-opponent computations. In sum, we provide strong, converging evidence of successful human cone modification leading to meaningful but incomplete rescue of visual function after life-long deprivation. More broadly, our findings suggest that the future of sensory augmentation technologies will depend not only on delivering new input to the nervous system, but also on understanding how neural circuits can effectively use it.

## Results

### Rescued cone signal

We first tested whether gene therapy successfully rescues functional cone signalling in children with achromatopsia. We developed cone-selective stimuli designed to be invisible to rod-mediated vision using silent substitution (Estévez & Spekreijse, 1982). This method generated colour pairs that activate rod photoreceptors identically despite differing spectral compositions (see Fig. 1 for visualisation). When presented as patterns (e.g., checkerboards) against a background, these stimuli are indiscriminable with rod-only vision but should be discriminable if there are functioning cones. This was validated through objective psychophysical testing in 17 untreated people with achromatopsia (Supplement 1).

**Figure 1.**
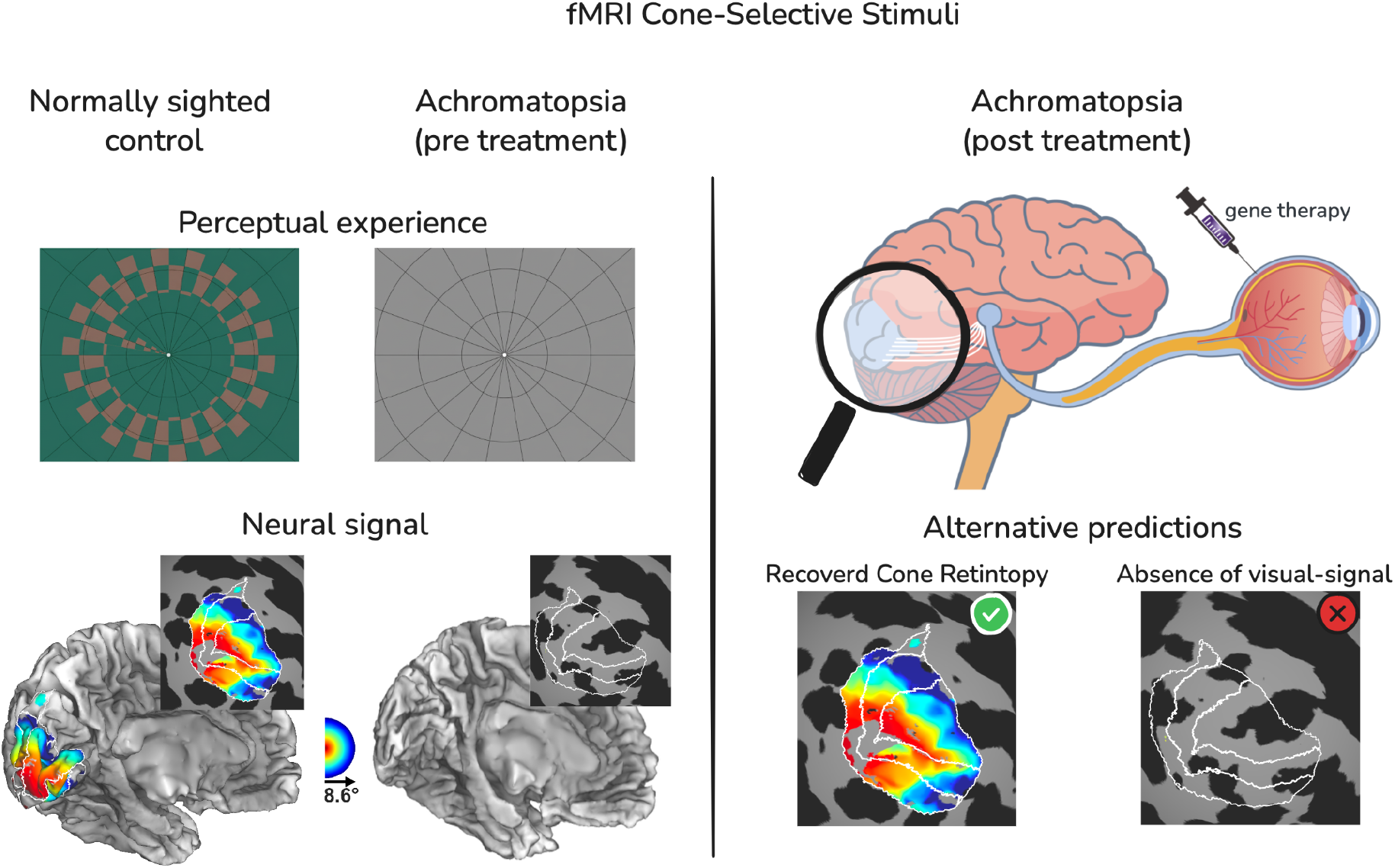
Experimental paradigm and predicted cortical outcomes of gene therapy. (**Left**) Cone-selective stimuli designed via silent substitution are perceived as contrasting patterns by normally sighted controls but remain invisible to untreated people with achromatopsia. Corresponding pRF eccentricity maps on inflated and flattened cortical surfaces illustrate the presence of a cone-mediated visual representation in controls and the absence of organised cone-mediated responses in people with achromatopsia prior to treatment. (**Right**) Alternative predictions for post-therapeutic outcomes; Successful cone-signal transmission should manifest as organised retinotopic maps, whereas a lack of recovery should result in a continued absence of visually driven cortical responses.

#### fMRI

Using fMRI, we tested whether genetic modification of non-functioning retinal cones can induce new visual signals in humans. We used population receptive field (pRF) mapping, a computational fMRI approach that systematically stimulates the retina and models how the visual cortex represents visual field locations. Combined with cone-selective stimuli, this approach allowed us to determine whether the retinal input was successfully restored and whether it engages the intact cortical architecture required for spatially precise, functional vision.

Figure 1 illustrates our approach and predictions. Before treatment, cone-selective stimulation should induce no organised response patterns in visual cortex as the cones are inactive, making the pattern invisible to the patients (Fig. 1). After treatment, successful genetic cone modification and signal transmission should manifest as stereotypical bilateral fMRI response patterns in visual cortex, as each eye projects to both cortical hemispheres. These should follow established retinotopic projections: eccentricity tuning shifts from central (red in Fig. 1) to peripheral (blue) locations from the occipital pole to anterior in V1-3 (Fig. 1), and polar angle tuning shifts from upper-to-lower field representations demarcating stereotypical boundaries between visual areas V1-3 (Supplement 2).

Prior to treatment, we found no cone-driven signalling in any of the patients. The proportion of vertices in visual cortex with above-threshold cone-selective signal was low (range: 0.01%-13.6%, at pRF *R² > 0.1,* see Supplement 3 for a full report) and lacked retinotopic structure (alignment-*R²* < 0), indicating spurious responses. Consistent with these neuroimaging results, no patient could perceptually detect the cone-selective stimuli during fMRI (Fig. 2A; validated via psychophysics, Supplement 1).

**Figure 2.**
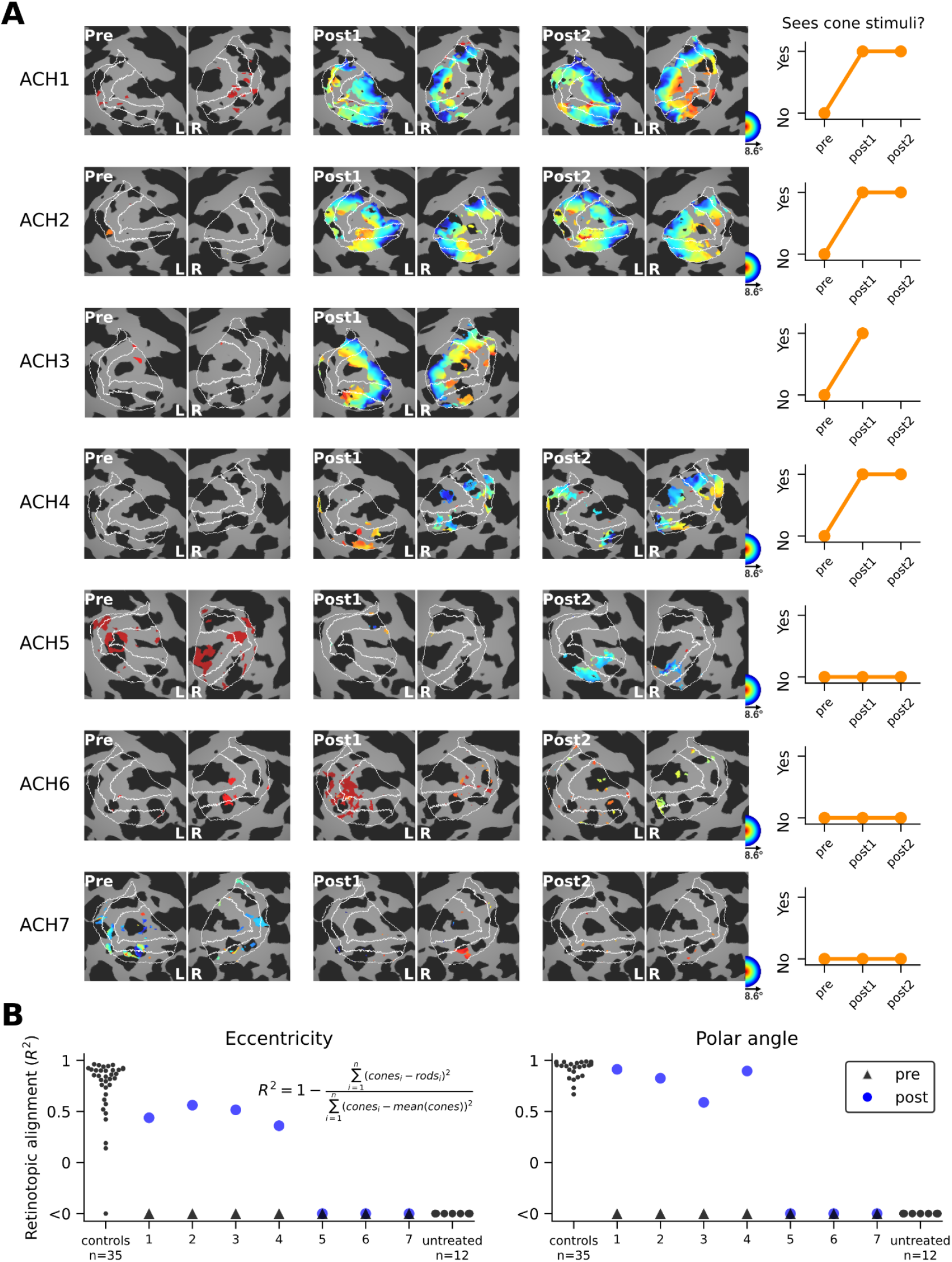
Rescued cone-signals in visual cortex. **(A)** Cone-selective visually-driven activity in V1-3 (pRF model fits R² > 0.1) in each patient before treatment and at their last two post-treatment follow-ups (11-35 months post-treatment). ACH3 did not return for a second post session. The maps are colour-coded based on eccentricity (i.e. distance in visual angles from the centre of fixation). There is a clear recovery of cone-driven retinotopic activity in ACH1-4 in two separate Post sessions compared to the Pre gene therapy measure. This neural change is paired with a shift in perception, where the cone-selective stimuli becomes visible to the patient (right column) **(B)** Clear presence of cone-mediated retinotopic structure in ACH1-4 is evident in the alignment (alignment-R²) between the eccentricity and polar angle values in the rod and cone map, which approaches the alignment distribution of 35 normally-sighted controls and is not present before treatment and in ACH5-7 post-treatment measures.

Following gene therapy, four of the seven treated patients (ACH1–4) exhibited marked and consistent cone-mediated cortical activity. The proportion of activated visual cortex reached 61% in ACH1, 56% in ACH2, 41.5% in ACH3 and 20% in ACH4 by their final post-treatment session. Crucially, these activations formed highly organised retinotopic patterns that are exceedingly unlikely to emerge by chance. Using each patient’s measured rod-mediated retinotopy as a topographic blueprint, we found that a spatial alignment metric of newly emerged cone-driven maps was comparable to the rod-cone alignment observed in normally-sighted controls (Fig. 2B, alignment-*R²* > 0.36).

These post-treatment cone maps were highly repeatable, demonstrating high correlations in eccentricity tuning across longitudinal sessions (Pearson’s r=0.70–0.88) and significant spatial overlap in activated cortical vertices (ranging from 27% to 72% overlap across sessions). This recovery of a spatially organised, stable cone signal represents a fundamental requirement for functional vision. Critically, this cortical recovery was accompanied by a perceptual breakthrough: patients ACH1–4, who were previously blind to the cone-selective stimuli, gained the ability to perceive them post-therapy - a significant measurable improvement in visual function - as confirmed by objective psychophysics (Supplement 1).

In contrast, patients ACH5–7 showed only spurious cone-mediated responses in visual cortex post-treatment (Fig. 2A). Only 1.5%–10% of voxels showed above-threshold responses, which lacked longitudinal consistency (ranging from 0 to 1.5% overlap) and demonstrated no spatial alignment with rod-mediated retinotopy (Fig. 2B). Consistent with these fMRI measures, these patients remained unable to perceive cone-selective stimuli, indicating rod-only vision (Supplement 1). Notably, robust rod-mediated retinotopic maps were successfully reconstructed for all seven patients across all sessions (Supplement 4), confirming that the absence of measurable cone signalling in ACH5-7 is not an artefact of insufficient data quality.

#### Behavioural evidence of cone-mediated perception rescue

The fMRI results are corroborated by independent objective psychophysical testing, which demonstrates that cone function rescue is restricted to the treated eye and persists for up to three years post-surgery. We employed a cone-contrast sensitivity task to quantify functional recovery. In this task, stimuli vary in their discriminability from easy (high cone contrast) to hard (low cone contrast). While initial levels are detectable via rod-mediated vision, the final six contrast levels are rod-isoluminant (i.e., invisible to rods; Farahbakhsh et al., 2022). We used a four-alternative forced-choice (4AFC) paradigm (Fig. 3A) with a staircase procedure, adjusting stimulus contrast based on performance, to define cone-mediated detection thresholds per patient. Performance ranges for rod-based vision were established using a cohort of 17 untreated people with achromatopsia (Fig. 3A) and meet predictions based on spectral characteristics of the stimuli (see Farahbakhsh et al., 2022 for more details).

**Figure 3.**
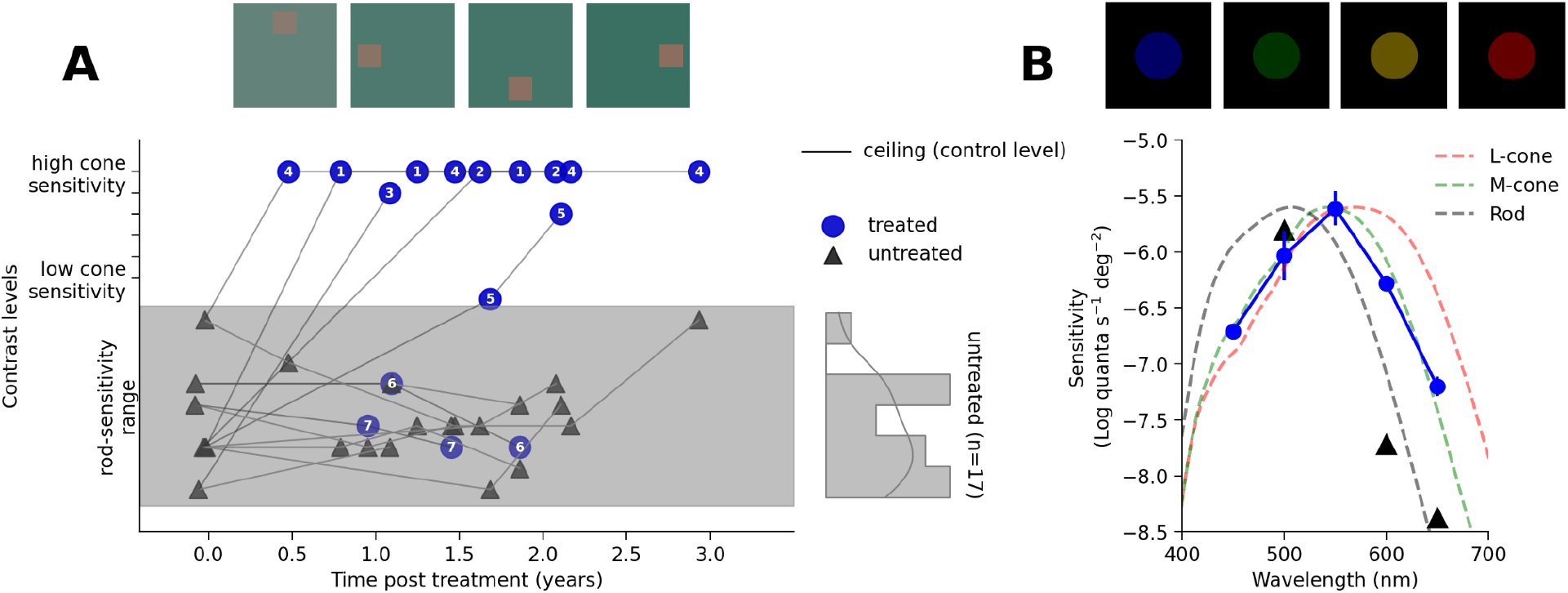
Behavioural evidence for recovery of cone-mediated perception. **(A)** Longitudinal measures from the cone-contrast sensitivity test. Top: Illustration of stimuli. In each trial participants indicated square location. Bottom: cone-contrast thresholds for treated (blue circles) and untreated (dark grey triangles) eyes over time. In the pre-treatment session the task was performed binocularly as both eyes were untreated. In all post sessions monocular data is plotted for each eye. Plotted rod-sensitivity range (grey shaded area) was established from a larger cohort of untreated achromats (n=17, right-hand histogram). ACH1-4 showed a significant gain in sensitivity close to or at control ceiling levels (horizontal line) selective to their treated eye. Non-responding participants (ACH6, ACH7) remained within the rod-mediated baseline range. **(B)** Spectral sensitivity results in ACH4. Top: Illustration of stimuli. In each trial participants adjusted target radiance until it was just visible. Bottom: Sensitivity (log quanta per second per degree squared) as a function of wavelength (nm). The untreated eye (triangles) aligns with the scotopic rod sensitivity curve (grey dashed curve). In the treated eye (blue circles), sensitivity is shifted, matching the M-cone tuning function (green dashed curve). Dashed lines have been vertically aligned to the peak of the treated eye. Error bars represent ±1 SEM.

All patients exhibiting cortical recovery (ACH1–4) demonstrate marked and sustained improvements in cone contrast sensitivity performance, reaching the ceiling levels observed in normally-sighted controls (Fig. 3A). A fifth patient (ACHM5) shows mild improvement, while ACHM6 and ACHM7 remain within the rod-performance range. Crucially, these gains are strictly specific to the treated eye while performance in the untreated eye remains within the baseline rod-range, ruling out systemic learning effects. This post-therapy benefit remains stable across all longitudinal follow-ups, with no observed decline up to three years post-surgery.

To further confirm the recovery is mechanistically cone-driven, we performed in-depth spectral sensitivity characterization of the genetically modified cones in ACH4. In this task patients view monochromatic targets of different wavelengths and adjust their radiance (amount of light) until the target is just visible (see Methods for more details). In the treated eye, we identify a definitive shift in peak sensitivity, away from a rod-dominated profile toward longer wavelengths, which closely matches the M-cone spectral tuning function (Stockman, 2019; Fig. 3B). This shift is not present in the untreated eye, confirming monocular restoration of cone-specific signalling due to treatment.

Robust convergence with fMRI and high-precision spectral sensitivity measures, positions the cone-contrast task (Fig. 3A) as a promising short, highly sensitive, child-friendly, and low-cost endpoint for assessing cone-signal restoration in future interventions.

#### Clinical predictors of cone rescue

We identified no systematic differences between responders and non-responders regarding age, dose, genotype, or OCT-derived markers of macular health (Table 1). Therefore, the factors that determine why a minority of patients show no therapeutic response on our measures remain unclear.

So far, our data provide definitive evidence that gene therapy can successfully restore signalling in cone photoreceptors that have remained dormant since birth, in at least four of seven patients (aged 8–15 years). Critically, these signals are retinotopically organised, longitudinally stable, and accessible to conscious perception, offering key building blocks for cone-mediated visual function gain.

The tests described above, were designed to maximise sensitivity to rescued cone signals by isolating cone-mediated input, reducing potential masking from dominant rod vision, and by minimising reliance on specialised post-photoreceptoral computations that may not be in place. Having established the presence of new cone-mediated retinal signalling we next examined which downstream pathways and visual functions can make use of them.

### Rescue of cone-specific functions

We next assess the functional impacts of the rescued cone signals on sight. We focus on visual domains where cone-mediated vision is most distinct from rod-mediated vision, to highlight benefits and bottlenecks of gene therapy for achromatopsia when treating beyond the 9th year of life. Specifically, we test whether genetically modified cones enable high temporal resolution (speed), recover foveal high-acuity vision, and support colour perception for the first time.

#### High-frequency temporal processing (speed)

Cones and their pathways are faster than rods and their pathways (Conner, 1982), due to phototransduction speed and downstream circuitry. Will the genetically modified cones support faster vision? We tested this in ACH4 using a critical flicker fusion task, where we showed a red (650nm) flickering target superimposed on a blue rod-saturating background, and identified how fast (high-frequency) the flicker needs to be for the target to appear static - the critical flicker frequency.

In controls, the highest detected flicker frequency increases with stimulus intensity (radiance), reaching ∼40 Hz (Fig. 4A). In ACH4, as intended, the task is highly challenging under rod-only vision in the untreated eye: the red target is largely undetectable, with flicker perception limited to ∼4 Hz at the highest radiance (Fig. 4A). By contrast, the treated eye detects targets at ∼100-fold lower radiances and detected flicker up to ∼30 Hz (Fig. 4A). This level of performance exceeds what can be achieved with rod-only vision.

**Figure 4.**
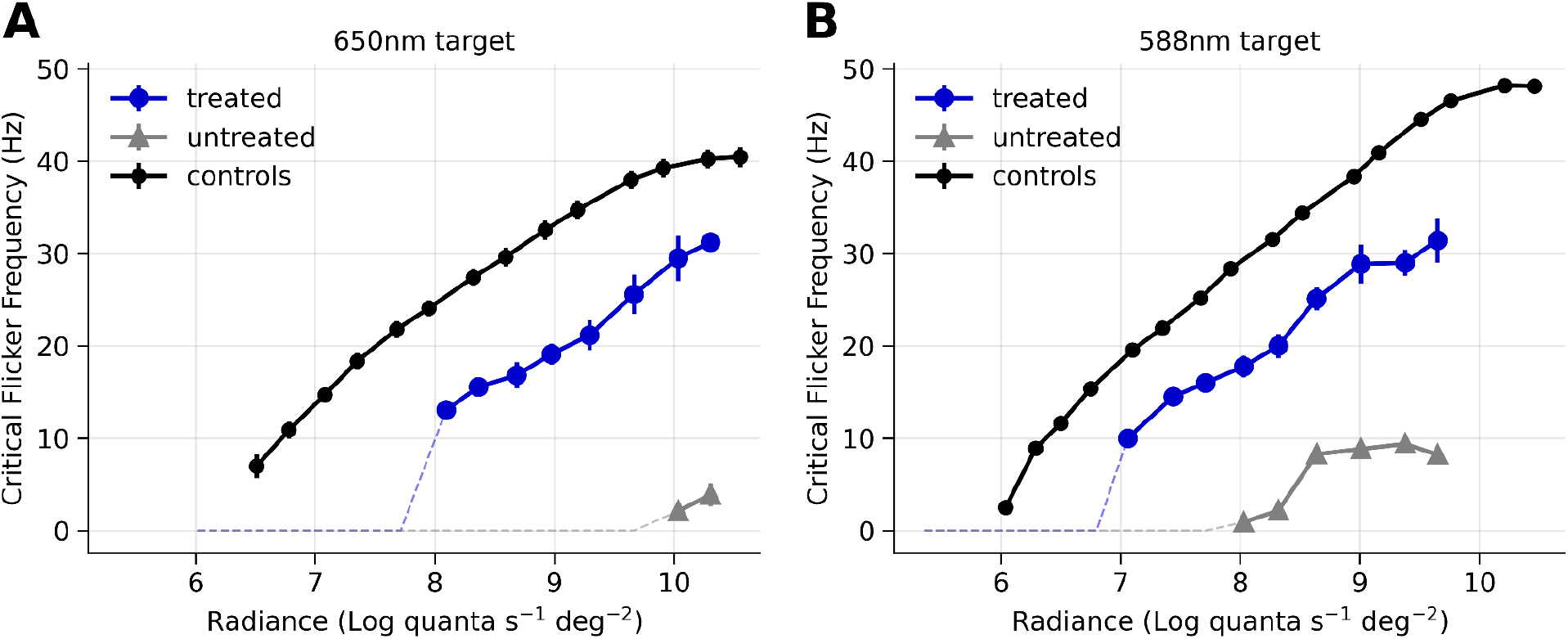
Recovery of high-speed temporal processing in ACH4. **(A)** Critical flicker fusion frequencies for a long-wavelength (650 nm) target. Temporal resolution was measured as a function of stimulus radiance (intensity). Normally sighted controls (black circles, n=12) show a characteristic increase in flicker frequency as radiance increases, reaching frequencies of ∼40 Hz. ACH4’s untreated eye (grey triangles) detected flicker only at very high radiance and very low temporal frequencies. In contrast, the treated eye (blue circles) shows a robust emergence of high-frequency flicker detection that approaches control levels (n=12). **(B)** Critical flicker fusion frequencies for a 588 nm target. A normally sighted control shows a characteristic increase in flicker frequency with light level, reaching almost 50 Hz. This is higher than for 650 nm as both L and M cones are sensitive to the 588 nm target. For the 588nm stimuli, the untreated eye (grey triangles) exhibit a response characteristic of rod-mediated vision, plateauing around 10 Hz. The treated eye (blue circles) demonstrates a marked increase in temporal resolution, shifting toward the performance profile of the healthy control. Dashed lines indicate the targets that were not visible at the tested radiance intensity. Error bars represent ±1 SEM

We replicated these results with a yellow target (588 nm; Fig. 4B). By comparing the change in sensitivity between the 650 and 588 nm targets, we can predict which photoreceptor(s) are likely detecting the flicker. Each type of photoreceptor has a different spectral sensitivity (Fig. 3B) and will therefore respond differently to the two wavelengths and generate a particular shift in temporal sensitivity. In the treated eye, the wavelength-dependent shift of the critical flicker frequency measures (∼0.81 log units) closely matches the prediction for L-cone sensitivity (0.79 log units), whereas the untreated eye shows a shift consistent with rod sensitivity (2.03 log units). While the spectral sensitivity measurements in Fig. 3B suggested a stronger M-cone contribution in ACH4, the flicker measurements point to a stronger L-cone contribution. This difference may reflect task-specific factors, including the retinal regions stimulated. Together, these complementary measures suggest that both L- and M-cone signals are present in ACH4’s treated eye

These psychophysical data demonstrate that genetically modified cones in ACH4 support high-frequency temporal processing, confirming that both the cone photoreceptors and sufficient downstream circuitry for fast cone-mediated vision remain available for rescue.

#### Foveal high-acuity vision

High-acuity vision, the ability to perceive fine spatial details, is mediated by the fovea, a small central retinal region densely packed with cone photoreceptors which, importantly, is completely devoid of rods, even in achromatopsia (Georgiou et al., 2019). As a result, unlike the rest of the retina, neural pathways supporting foveal input receive no rod input so are completely light-deprived from birth. This raises a critical question: can high-acuity foveal vision be restored after more than a decade of sensory deprivation?

Before treatment, optical coherence tomography (OCT) confirmed that the gross retinal architecture required for foveal function was preserved across all treated patients (Fig. 5A). In particular, patients showed a largely intact ellipsoid zone and a substantial outer nuclear layer, indicating the continued presence of cone photoreceptors (Table 1).

**Figure 5.**
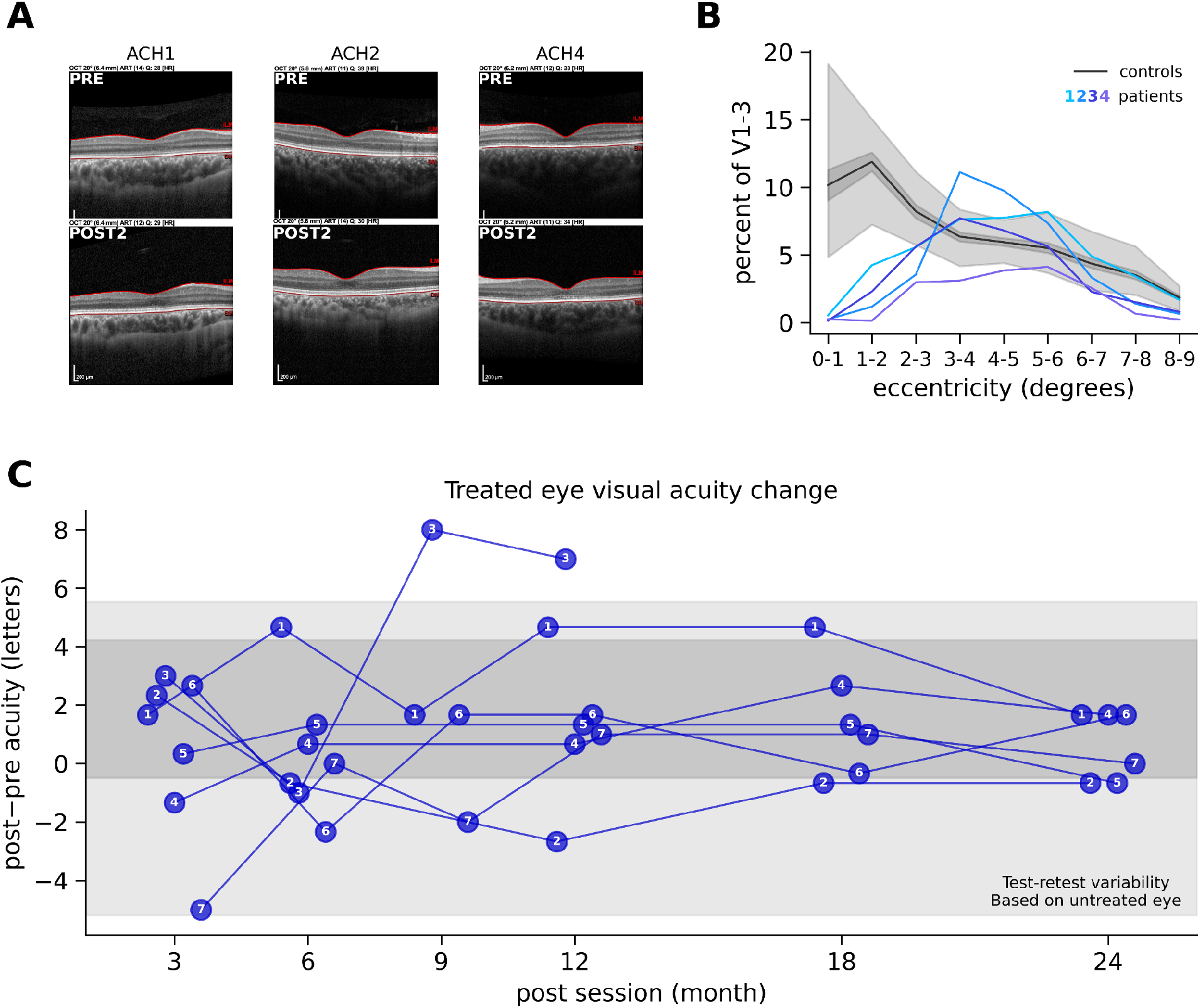
Foveal vision recovery. **(A)** Spectral-domain optical coherence tomography (SD-OCT) cross-sections through the fovea of responding participants (ACH1, ACH2, ACH4). Retinal layering and foveal architecture remained stable between baseline (PRE) and approximately two years post-treatment (POST2), showing preserved foveal infrastructure (PRE) and no significant structural degeneration or adverse effects from the surgical intervention (POST). (**B)** Eccentricity distribution of cone-selective cortical signals. Proportion of pRFs as a function of eccentricity (in degrees) in V1–V3 for controls (n=39, black) and the four responding participants (blue). In controls, pRF density is highest in the central 0−1°. In treated patients, rescued cone signals are significantly biased toward the periphery, with a marked deficit in foveal representation (0-2°). Shaded areas represent ±1 SEM (dark grey) and 95% prediction intervals (light grey). **(C)** Longitudinal visual acuity change. Difference in letters correctly identified (Post − Pre) over 24 months for the treated eye. Most participants remained within the range of test-retest variability established by untreated eyes of the same individuals across all follow-up sessions (mean ±1.96 SD, dark grey; full range, light grey).

Despite this preserved structure, post-treatment visual acuity showed no consistent improvement on ETDRS letter charts, a primary endpoint in many ocular therapy trials. Only one younger participant (ACH3) exhibited gains exceeding the test–retest range of all patients’ untreated eye measures (Fig. 5C). Because achieving high acuity depends specifically on foveal cones and their post-receptoral pathways, this limited improvement suggests that high-acuity foveal vision remained constrained.

To test for evidence of foveal cone signal rescue, we examined cone-driven visual field representations in visual cortex (V1-3) as assessed by the pRF mapping fMRI (further analysing data from Fig. 2). Patients show limited cone-driven signals from the central fovea (0–1°; Fig. 5B), which in normally sighted individuals innervate a large cortical territory. This may indicate limited recovery of foveal vision, although some foveal signals may be present but obscured by fixation instability.

From a neuroplasticity perspective foveal rescue in achromatopsia may be particularly difficult beyond early sensitive periods; unlike in the peripheral retina, the rod-free fovea never receives alternative local light-driven input to maintain downstream circuitry in the absence of cone function (see Discussion). Defining where bottlenecks arise from photoreceptor to cortex, and when they become limiting, may be key for optimising treatments for acuity gain.

#### Colour vision and cone-rod luminance integration

After establishing that patients can perceive rescued cone signals, and use them to detect stimuli that were previously invisible to them (Fig. 3 and 4), we next asked what perceptual information these signals convey. Full cone rescue is expected to alter the appearance of objects in both luminance (brightness) and colour. Do the cone signals only integrate into existing rod-based luminance pathways, resulting in achromatic vision with expanded photopic sensitivity, or do they enable the perception of colour for the first time?

Two patients with clear treatment effects (ACH2 and ACH4) completed a Rayleigh matching task: a sensitive test that characterises how colour and luminance information are used to generate perception (Birch, 1998; Kohl et al., 1993). The apparatus (Fig 6A) is made of two half-circles, with the top half containing a mixture of red and green monochromatic lights (mixture colour), and the bottom containing a yellow monochromatic light (target colour). Participants are asked to adjust the stimuli until the halves appear identical.

**Figure 6.**
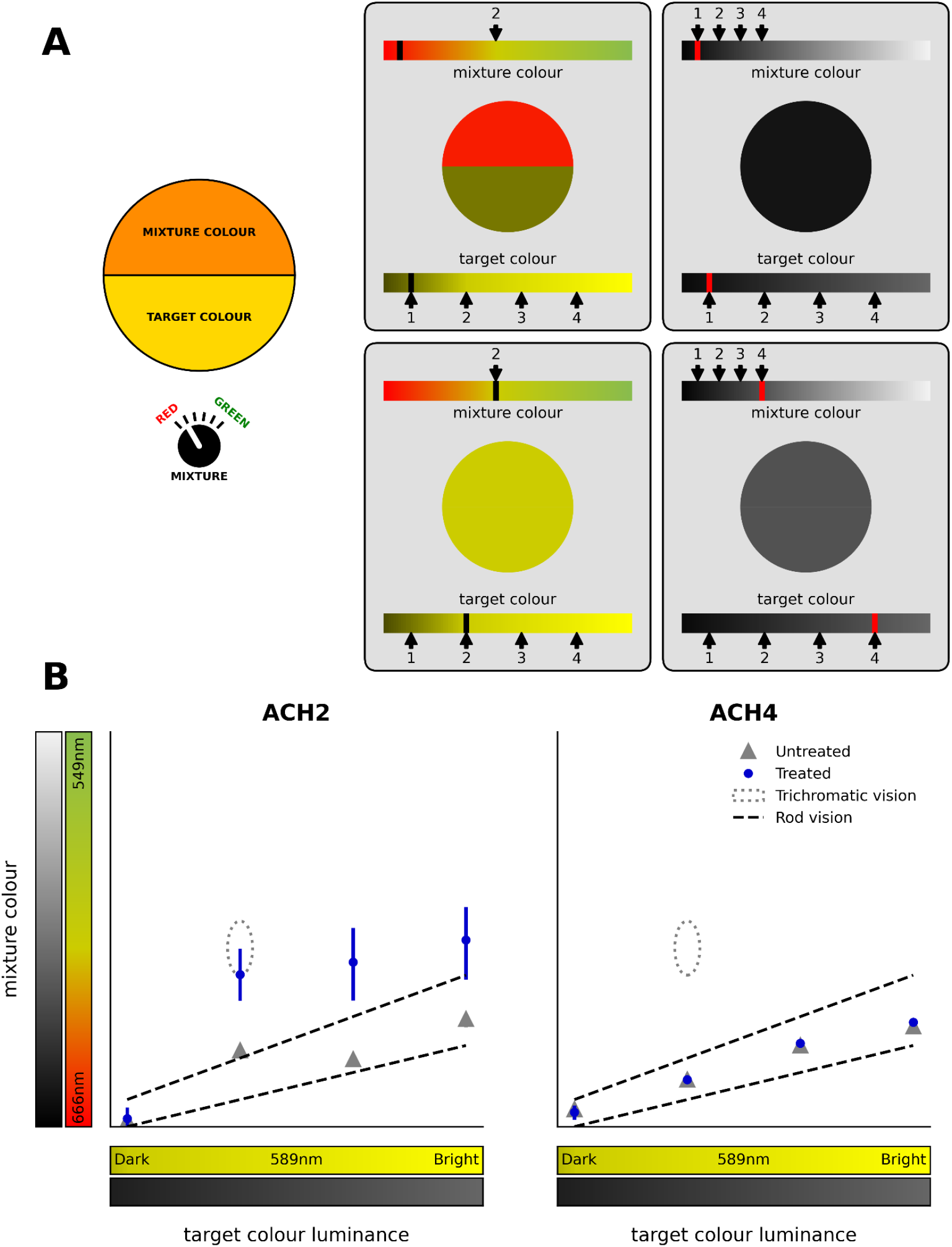
Colour vision and luminance perception. Rayleigh matches results using OCULUS HMC Anomaloscope. **(A)** Participants adjusted the red-green mixture ratio (mixture colour, top) to match a monochromatic 589 nm yellow target light (target colour, bottom) at 4 luminance levels (marked with arrows at the bottom colourbars). For normal trichromatic colour vision (first column), a match is impossible for targets 1,3,4 (target 1 is shown as an example). For target 2 a match is possible at a specific mixture level, this match is indicated with an arrow on the top colour bar and is represented in B as a dotted circle. For achromatic (rod-mediated) observers (second column), matches are possible for any yellow luminance as the two fields are equated only based on their perceived brightness. Examples for target 1 and 4 matches are plotted. Rod-based matches can be predicted based on the rod spectral sensitivity and are plotted in B as the “rod vision range”. **(B)** Red-green mixture settings are plotted against fixed yellow target luminance levels for treated (blue circles) and untreated (grey triangles) eyes. Settings made with untreated eyes across both participants (ACH2, ACH4) fell along the predicted rod-mediated luminance-match line. In their treated eyes, neither showed typical trichromatic matching. ACH2 showed a shift in luminance matching away from the rod vision range. ACH4’s matches remained within the rod vision range however they reported the two halves never fully perceptually matched. Error bars represent 95% confidence intervals. Variability in responses of the untreated eye of ACH2 and in both eyes of ACH4 was so small that the intervals are smaller than the markers themselves.

In normal colour vision (trichromatic vision), a match is only possible for a precise ratio of the red and green light mixture at a specific yellow luminance (visualised in Fig. 6A and plotted as a dotted ellipse in Fig. 6B). This is because we do not perceive colour directly - colour perception results from opponent circuits that compare signals from cones with different spectral sensitivities. For people with normal colour vision, the red and green mixture must produce the same L- and M-cone excitations as the pure yellow light so that they match in both cone-opponent and luminance postreceptoral pathways.

Since people with achromatopsia have only rods, they are colour blind and able to see variations in luminance but not in colour. The Rayleigh matches can therefore be made between any red–green mixture and yellow purely by matching the luminance of the target and the match (right, Fig. 6A; dashed lines, Fig. 6B).

If chromatic (cone-opponent) vision is rescued, matches on this task should be constrained to a small range of the mixture–luminance space in Fig. 6B. In contrast, if vision remains achromatic (luminance-based) matches should fall on a line spanning multiple yellow luminances. Moreover, the slope and the position of this line should provide information about which photoreceptor signals (rod or cone) that contribute to luminance perception.

With untreated eyes, as expected, both patients show rod-like matching. In treated eyes, the patients’ responses diverge. ACH2 shows no evidence of colour opponency, accepting matches across all yellow targets consistent with luminance-based matching. However, matches from the treated eye are shifted away from the rod monochromatic line, suggesting combined rod and cone contributions to luminance perception. This result aligns with ACH2’s subjective report that vision appears “brighter and slightly better” through the treated eye, which was previously their worse-seeing eye.

In contrast, ACH4’s matches remain within the rod vision range (Fig. 6B) despite clear cone rescue in all previous measures, suggesting the continued dominance of rod-mediated luminance perception with no evidence of colour opponency. Intriguingly, however, ACH4 consistently reported a subjective difference in their treated eye: the red–green mixture never appeared to match the yellow target perfectly due to a “subtle shimmer” present only in the mixture field. In daily vision, ACH4 also finds that reds and other warm colours have acquired a distinctive, “neon-like” quality. This experience has also been reported by patients from a different cohort receiving gene therapies for achromatopsia (McKyton et al., 2023).

In line with these subjective experiences, ACH4 showed objective evidence of some colour perception across two independent tasks: the Low Vision Cambridge Colour Test and a custom hue/saturation colour arrangement test (Supplementary 5). Only with their treated eye, they successfully discriminated colour targets from achromatic luminance distractors. Given the absence of canonical cone opponency in the Rayleigh matches, these effects are unlikely to reflect typical colour vision. Instead, they likely depend on non-canonical cues, such as luminance jitter from spatial inhomogeneities in the cone mosaic during small eye movements, or temporal differences between rod- and cone-mediated signals within achromatic pathways. Such processes could be consistent with the shimmery percept of colour. By contrast, ACH2 showed no colour discrimination on these tasks, consistent with their lack of detectable cone-opponency.

Together, these results point towards mixed neuroplasticity capacity. When treated later in life (>8 years in this study), rescued cone signals can still alter luminance perception and introduce novel colour cues. However, the recovery of classical cone-opponent computations may require earlier intervention.

## Discussion

We provide strong converging evidence that gene therapy can restore functional cone-mediated input in the human visual system after many years of congenital deprivation. In four of seven children with *CNGA3*-associated achromatopsia treated between 8 and 15 years of age, genetically modified cones generated robust retinotopically organised responses in early visual cortex that were absent before treatment. These newly established signals enabled perception of stimuli that had previously been invisible, were specific to the treated eye, and remained stable across all follow-up sessions up to 3 years (Figs 2, 3 & 4).

Achieving this level of rescue after years of total cone inactivity is striking. Unlike previous ocular gene therapies in humans, which largely restored retinal function that had once been present, our results show that cone photoreceptors, silent from birth, can be activated, introducing an entirely new signal to the brain. Critically, we found that downstream pathways are sufficiently developed and preserved to support the transmission and conscious perception of these novel visual signals. This marks an important step for sensory rescue and augmentation, showing that newly restored input can support meaningful cortical and perceptual function even after prolonged congenital deprivation.

The consistent evidence for cone-mediated rescue presented here, stands in contrast to prior reports. While several achromatopsia gene therapy trials have established the safety and feasibility of retinal gene delivery, standard clinical endpoints, such as visual acuity, contrast sensitivity, and colour tests, showed only modest and variable evidence of cone rescue across patients, sessions, and measures (Fischer et al., 2020; Michaelides et al., 2023; Reichel et al., 2022). We propose that standard visual function endpoints can obscure successful rescue for two reasons. First, performance in many endpoints may implicitly rely on multiple downstream computations that may not be readily available for the new signals after lifelong deprivation. Second, most standard assessments do not isolate cone function. When new cone signals are introduced alongside pre-existing rod signals, their contribution may initially be relatively weak or down-weighted. In tasks that recruit both rod- and cone-mediated processing, these cone contributions may therefore be masked or diluted.

By using cone-selective tests designed to maximise sensitivity to rescued cone signals while minimising reliance on specialised downstream processing, we show that rescue can be detected even when full functional recovery remains incomplete. This distinction is critical. Modest functional gains do not necessarily indicate failure of the treatment at the photoreceptor level, but may instead reflect downstream bottlenecks that could potentially be addressed. Extensive work in amblyopia and congenital cataract shows that earlier intervention and targeted rehabilitation are key to achieving the best functional outcomes (Holmes et al., 2011; Holmes & Levi, 2018; Maurer, 2017). Further advances in gene therapy technologies are also likely to improve outcomes, but full functional recovery may ultimately depend on addressing neuroplasticity constraints.

Beyond demonstrating the emergence of new cone signals and their successful relay to visual cortex, our results show that genetic cone rescue in achromatopsia can support meaningful visual function. Comprehensive psychophysical testing revealed faster temporal processing, a function strongly associated with cone-mediated pathways. In one individual (ACH2), we observed a robust shift in luminance perception, indicating that newly restored cone signals are being integrated into rod-mediated vision. In a second individual (ACH4) we found evidence for a non-canonical colour-related cue superimposed on rod-based luminance perception, consistent with the patient’s subjective reports.

At the same time, recovery was selective. We found that evidence for improvement in foveal acuity and colour processing was limited, suggesting that some specialised functions remain constrained despite the presence of new cone signals. What makes some visual functions more amenable to rescue later in life than others? One possible mechanistic explanation is that recovered functions draw on circuitry maintained by rod-driven activity. Rod and cone pathways share portions of retinal and post-retinal processing (Kolb et al., 1995; Sharpe & Stockman, 1999; Solomon, 2021). In the retina, rod signals can propagate into neighbouring cone circuitry through gap junctions or amacrine-cell pathways, potentially keeping the cones and their direct downstream circuitry active despite absent cone phototransduction. This propagation of rod input into the cones might contribute to the relative preservation of the normal structure of the retina in achromats. Stockman et al., (1995) argued that the existence of fast rod signals in achromatopsia, which in normal observers were thought to be transmitted via rod-cone gap junctions, implied the existence of functioning cone inner segments in achromatopsia, which was consistent with them having normal inner nuclear and ganglion cell layers (Glickstein & Heath, 1975).

Further downstream, from the level of the ganglion cells onwards, rods and cones also converge onto shared pathways. The magnocellular circuitry that supports achromatic fast signalling is shared across both photoreceptor classes and is most likely maintained by the rods. Treated cones may therefore be able to engage pre-maintained circuits, providing a possible explanation for the recovery of temporal processing and changes in luminance perception.

By contrast, functions that showed less recovery in our data may rely on circuitry without such a rod-driven “lifeline”. The central fovea which supports high-acuity vision is rod-free and therefore receives little or no surrogate rod input during years of cone inactivity. Likewise, canonical colour vision depends on cone-opponent computations that cannot be established by rod signalling. Although rod signals can propagate into neighbouring cones through gap junctions, they will provide identical input to L- and M-cones. This is unlikely to support experience-dependent refinement and maintenance of cone-opponent midget-parvocellular pathways that rely on differential L/M signalling for red-green colour vision. Therefore, parvocellular cone circuitry, which supports both high acuity and chromatic vision, is likely either not to develop or to develop in an atypical way, creating a bottleneck even when cone signals are rescued. A key question for future studies is whether post-treatment experience can subsequently engage and refine cone-opponent circuitry, and how this depends on age when the treatment is administered.

Future work should also dissociate whether limited foveal recovery reflects incomplete photoreceptor rescue or downstream circuit constraints, for example using stimulus-evoked adaptive optics to measure photoreceptor activity directly (Harmening & Sincich, 2019; Jonnal, 2021). Notably, a recent clinical trial report found evidence of activity in ∼25% of foveal cones in the treated eyes of patients with achromatopsia, measured as wave-guided reflectance using adaptive optics (Jiang et al., 2024). This change was only present in 2 of the 18 treated patients, who were among the youngest in the study (<10 years). This shows that foveal photoreceptor rescue is possible, but may depend on early treatment. The promise of early intervention is supported by recent *AIPL1* gene therapy in severe Leber Congenital Amaurosis (LCA; Michaelides et al., 2025), where treatment delivery before the age of 3 years produced large gains in acuity, colour vision and self-reported daily function.

For congenital cone disorders, the implications of our study are clear: gene therapy can rescue photoreceptors dormant from birth, and while specialised cone functions may only partially recover after long-term deprivation, there is strong potential for fuller rescue through earlier intervention, rehabilitation and therapeutic optimisation. For other retinal disorders, and for sensory augmentation interventions more broadly, the relevant neuroplasticity constraints may differ, and so too will the solutions. The broader principle, however, remains the same: delivering new signals to the brain is only the first step. The success of augmentation depends on understanding how those signals can be integrated to improve perceptual function. Achieving this will require approaches that trace signals through the nervous system, reveal function-specific plasticity constraints, and guide interventions that unlock the maximal capacity for recovery.

## Methods

### Participants

This study’s primary cohort consists of seven patients with achromatopsia who received gene therapy in one eye. All participants had genetically confirmed achromatopsia, with bi-allelic disease-causing variants in *CNGA3;* additional data is provided in Table 1. They also met MRI safety inclusion criteria and had no other known neurological disorders.

In the cone-selective psychophysics task an additional ten participants with variants in either *CNGA3* or *CNGB3* were tested to generate a baseline performance benchmark for the task. All participants met MRI safety inclusion criteria and had no other known neurological disorders. Data collection had ethics approval (separate from clinical trials) from the national ethics committee for patients (REC reference: 12/LO/1196; IRAS code: 106506). OCT and visual acuity data were collected under the CNGA3-ACHM gene therapy phase I/II clinical trial (NCT03758404) ethics approval (REC reference: 19/LO/0041)

Most participants completed one pre-treatment and two post-treatment fMRI sessions. Post-treatment sessions occurred at approximately 12 months (range: 6–20) and 20 months (range: 15–25) after intervention. Deviations from this schedule included ACH3 (one post-treatment session), ACH1 (one additional session), and ACH4 (two additional sessions). For ACH1 and ACH4, the final session is plotted as ‘Post 2’ using monocular data from the treated eye; binocular maps from the same sessions yielded similar or slightly attenuated patterns.

Forty normally-sighted controls took part in the fMRI portion of the study. Sixteen children: mean age = 10.75, range = 6–15 years; 24 adults: mean age = 25.79, range = 18–34 years). One control participant did not complete the cone-selective stimuli condition and four did not complete the rod-selective condition. Data collection had ethics approval from the UCL ethics committee (#4846/001).

### MRI

#### MRI Acquisition protocols

Participants were scanned in a Siemens Avanto 1.5T MRI scanner with a 30-channel coil (a 32-channel coil customised to remove view obstructions). A high-resolution structural scan was acquired using a T1-weighted 3D MPRAGE (1 mm^3^ voxel size, Bandwidth = 190 Hz/Px, 176 partitions, partition TR = 2730, TR = 8.4ms, TE = 3.57, effective T1 = 1000 ms, flip angle = 7 degrees). Functional T2*-weighted multiband 2D echo-planar images were acquired using a multi-band sequence with 2.3 mm isotropic voxels (TR = 1000 ms, TE = 55 ms, volumes = 348, flip angle = 75°, MB acceleration factor = 4, Bandwidth = 1628 Hz/Px).

#### fMRI stimuli and apparatus

Stimuli were presented on an MR-compatible LCD display (BOLDscreen 24, Cambridge Research Systems Ltd., UK; 51 x 32 cm; 1920 x 1200 pixels) viewed through a mirror in the scanner at 105 cm distance. Participants were lying supine in the scanner, with fixation stability recorded where possible via a mirror, with an Eyelink 1000 Plus (SR Research, Ottawa, ON) at the back of the bore. Built-in gaze calibration was not possible in people with achromatopsia due to nystagmus so the camera was calibrated in advance on a healthy eye. Before every other run, patients fixated on a 5-point custom calibration which allowed us to calibrate gaze measures post-hoc (Tailor et al., 2021). Stimuli were presented using MATLAB (R2016b, MathWorks, Natick, MA, USA), via the Psychophysics Toolbox 3 (Kleiner et al., 2007). pRF mapping stimuli comprised a simultaneous rotating ring and contracting/expanding wedge embedded within a contrast-reversing checkerboard with 2Hz reversal rate. Per run, the ring expanded/contracted for 6 cycles (48 secs/cycle) with logarithmic eccentricity scaling (van Dijk et al., 2016). The wedge (20° angle) rotated clockwise/anticlockwise for 8 cycles (36 sec/cycle). 20-second fixation baselines were embedded at the start, at the mid-point, and end of the run (total run duration 348 secs). The stimuli covered a maximum eccentricity of 8.6°, and moved to a new position each 1-second TR. The stimulus was overlaid with a small white central fixation dot (0.2° VA radius) and a black radial grid to encourage stable fixation. To keep participants engaged, they played a rewarded “kitten rescue mission game” in which they pressed a button when detecting a fixation target luminance change.

Cone-selective stimuli: To selectively measure cone-mediated signal processing in fMRI, we created a pair of colours (chromatic pairs) carefully calibrated to induce different levels of activation in cone photoreceptors but identical levels of activation in rods (see Fig. 1), thus making them indiscriminable to rod photoreceptors (silent substitution) (Estévez & Spekreijse, 1982; Spitschan & Woelders, 2018). For an in-depth description of stimulus generation and validation see Farahbakhsh et al., 2022. In brief, the cone-selective colour pair was generated by only varying the red and green channels of the RGB screen, keeping the blue constant. This shifts the stimulus variations to longer wavelengths, which the rods are less sensitive to, thereby reducing potential errors in rod equating. Neutral density filters were used to present these stimuli in the mesopic/low photopic light range (max luminance: 0.8 cd/m^2^), to keep viewing tolerable for this photosensitive population.

Rod-selective stimuli: To selectively measure rod-mediated signals we generated a single chromatic pair that kept L and M cone contrast constant (i.e., invisible to L and M cones), whilst inducing a contrast response in rods. Since it is only possible to simultaneously silence two photoreceptor types with 3 colour channels, the resulting stimulus was not controlled for S-cone contribution. To minimise the effects of the S-cone response, we presented these stimuli at very low light level (max luminance: 0.02 cd/m^2^). Prior to the rod-selective scan, participants had 15-min of dark adaptation while they listened to a story and structural scans were acquired.

#### MRI pre-processing

Data were preprocessed using *fMRIPrep* 21.0.2 (Esteban, Blair, et al., 2018; Esteban, Markiewicz, et al., 2018; RRID:SCR_016216), which is based on *Nipype* 1.6.1 ((K. Gorgolewski et al., 2011; K. J. Gorgolewski et al., 2018; RRID:SCR_002502).

#### fmriprep structural data pre-processing

The T1-weighted (T1w) image was corrected for intensity non-uniformity (INU) with N4BiasFieldCorrection (Tustison et al., 2010), distributed with ANTs 2.3.3 (Avants et al., 2008; RRID:SCR_004757), and used as T1w-reference throughout the workflow. The T1w-reference was then skull-stripped with a *Nipype* implementation of the antsBrainExtraction.sh workflow (from ANTs), using OASIS30ANTs as target template. Brain tissue segmentation of cerebrospinal fluid (CSF), white-matter (WM) and gray-matter (GM) was performed on the brain-extracted T1w using fast (FSL 6.0.5.1:57b01774, RRID:SCR_002823, Zhang et al., 2001). Brain surfaces were reconstructed using recon-all (FreeSurfer 6.0.1, RRID:SCR_001847, Dale et al., 1999), and the brain mask estimated previously was refined with a custom variation of the method to reconcile ANTs-derived and FreeSurfer-derived segmentations of the cortical gray-matter of Mindboggle (RRID:SCR_002438, Klein et al., 2017). Volume-based spatial normalization to one standard space (MNI152NLin2009cAsym) was performed through nonlinear registration with antsRegistration (ANTs 2.3.3), using brain-extracted versions of both T1w reference and the T1w template. The following template was selected for spatial normalization: *ICBM 152 Nonlinear Asymmetrical template version 2009c* [Fonov et al., 2009, RRID:SCR_008796; TemplateFlow ID: MNI152NLin2009cAsym]. Many internal operations of *fMRIPrep* use *Nilearn* 0.8.1 (Abraham et al., 2014, RRID:SCR_001362), mostly within the functional processing workflow.

#### fmriprep functional data pre-processing

For each BOLD run found per subject (across all tasks), the following preprocessing was performed. First, a reference volume and its skull-stripped version were generated by aligning and averaging 1 single-band references (SBRefs). Head-motion parameters with respect to the BOLD reference (transformation matrices, and six corresponding rotation and translation parameters) are estimated before any spatiotemporal filtering using mcflirt (FSL 6.0.5.1:57b01774, Jenkinson et al., 2002). The BOLD time-series were resampled onto their original, native space by applying the transforms to correct for head-motion. These resampled BOLD time-series will be referred to as *preprocessed BOLD in original space*, or just *preprocessed BOLD*. The BOLD reference was then co-registered to the T1w reference using bbregister (FreeSurfer) which implements boundary-based registration Greve & Fischl, 2009). Co-registration was configured with six degrees of freedom. First, a reference volume and its skull-stripped version were generated using a custom methodology of *fMRIPrep*. Several confounding time-series were calculated based on the *preprocessed BOLD*: framewise displacement (FD), DVARS and three region-wise global signals. FD was computed using two formulations following Power (absolute sum of relative motions, Power et al., 2014) and Jenkinson (relative root mean square displacement between affines, Jenkinson et al., 2002). FD and DVARS are calculated for each functional run, both using their implementations in *Nipype* (following the definitions by Power et al., 2014). The three global signals are extracted within the CSF, the WM, and the whole-brain masks. Additionally, a set of physiological regressors were extracted to allow for component-based noise correction (*CompCor*, Behzadi et al., 2007). Principal components are estimated after high-pass filtering the *preprocessed BOLD* time-series (using a discrete cosine filter with 128s cut-off) for the two *CompCor* variants: temporal (tCompCor) and anatomical (aCompCor). tCompCor components are then calculated from the top 2% variable voxels within the brain mask. For aCompCor, three probabilistic masks (CSF, WM and combined CSF+WM) are generated in anatomical space. The implementation differs from that of Behzadi et al. in that instead of eroding the masks by 2 pixels on BOLD space, the aCompCor masks are subtracted a mask of pixels that likely contain a volume fraction of GM. This mask is obtained by dilating a GM mask extracted from the FreeSurfer’s *aseg* segmentation, and it ensures components are not extracted from voxels containing a minimal fraction of GM. Finally, these masks are resampled into BOLD space and binarized by thresholding at 0.99 (as in the original implementation). Components are also calculated separately within the WM and CSF masks. For each CompCor decomposition, the *k* components with the largest singular values are retained, such that the retained components’ time series are sufficient to explain 50 percent of variance across the nuisance mask (CSF, WM, combined, or temporal). The remaining components are dropped from consideration. The head-motion estimates calculated in the correction step were also placed within the corresponding confounds file. The confound time series derived from head motion estimates and global signals were expanded with the inclusion of temporal derivatives and quadratic terms for each (Satterthwaite et al., 2013). Frames that exceeded a threshold of 0.9 mm FD or 1.5 standardised DVARS were annotated as motion outliers. The BOLD time-series were resampled onto the following surfaces (FreeSurfer reconstruction nomenclature): *fsnative*. All resamplings can be performed with *a single interpolation step* by composing all the pertinent transformations (i.e., head-motion transform matrices, susceptibility distortion correction when available, and co-registrations to anatomical and output spaces). Surface resampling was performed using mri_vol2surf (FreeSurfer).

#### Additional pre-processing

The first 4 volumes of each run were discarded for signal stabilization. Bold data in fsnative space was smoothed along the cortical surface (Gaussian kernel FWHM = 3 mm). For each run, a denoising procedure was further applied on the data by inputting nuisance regressors into a PCA and regressing out the top 5 components. The nuisance regressors derived from fMRIprep outputs and include: Six head motion regressors, FD and DVARS, CSF mean signal, white matter mean signal, the first five anatomical CompCor regressors (csf+wm combined), and the first five temporal CompCor regressors. Then data and regressors were temporally filtered with a Savitzky-Golay filter (3rd order, 347 seconds in length) and zscored. The time series from the two identical task runs were then averaged to increase signal to noise ratio.

#### MRI analysis

pRF modelling was carried out with a custom Python package, prfpy (Aqil & Knapen, 2023). To model population receptive fields, we used a two-dimensional isotropic Gaussian function (Dumoulin & Wandell, 2008), with x,y representing the preferred retinotopic position, spatial standard deviation sigma (σ) representing pRF size as well as the baseline of the timeseries and the signal amplitude beta (b). At every cortical location, the parameters of each model were optimized by minimizing the residual sum of squares between the model prediction and the measured BOLD signal. Fitting began with a grid search for the best model parameters (pRF position and size) followed by an iterative optimization. In each participant, in each condition, vertices with an R^2^ < 0.1 (variance explained by the pRF model), a negative beta, or a centre outside the stimulated radius (8.6°) were excluded from further analyses.

An anatomically defined retinotopy atlas was applied to the inflated cortical surface for each participant in order to delineate the borders of V1, V2 and V3 (Neuropythy v0.12.9; Benson & Winawer, 2018). Individual participant retinotopy maps were visualised on a flattened mesh of the occipital cortex.

For each participant, we quantify the structure of the cone retinotopy map using their own rod retinotopy map as a model. We calculate goodness-of-fit by dividing the sum of the squares of the residuals between the eccentricity and polar angle values across the rod and cone maps (SSR) by the total sum of squares (SST) (Polar angle distances were calculated as circular differences). This tests for full pointwise identity between the maps:

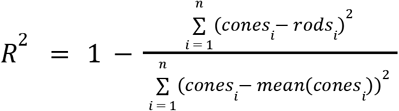

The closer to 1 the better the alignment is between the two maps. An R^2^<0 indicates that the rods map provides a worse prediction of the cone map than a null model that predicts each vertex the mean of the cone map.

### Screen calibration for silent substitution

To map between the screen and photoreceptor activations we computed transformation matrices to convert changes in the LCD screen’s red, green and blue (RGB) channel voltages, into changes in L-cone, M-cone and rod photoreceptor stimulation, using the RGB spectral output and the standard observer sensitivity functions for rod and cone photoreceptors (Stockman & Sharpe, 2000; Wyszecki & Stiles, 1982). Spectral output was measured with a Spectrascan Spectroradiometer, PR-655, PhotoResearch Inc., corrected for distortions from the fMRI mirror and neutral density filters. This allowed us to calculate the change in RGB voltage required to independently increment or decrement L- and M-cone or rod photoreceptor activity by prespecified proportions with respect to a baseline RGB value (mid-grey).

### Cone-selective psychophysics

A cone-contrast sensitivity test was used to assess cone signal functional recovery objectively; participants were asked to locate a square (3° of visual angle), presented at 6° eccentricity to the left, right, above or below a central marker (0.4° VA) in a 4 Alternative Forced Choice (4AFC) task. Participants viewed the display binocularly, were not required to fixate and had unlimited time to search. Each 4AFC 1-up/1-down adaptive staircase continued until at least 14 reversals (mean n trials = 27) or >8 correct responses had occurred at the lowest cone contrast (ceiling). Twenty-one contrast levels were generated using silent-substitution as equidistant steps between the highest and lowest steps achievable in our setup. Unfortunately due to a technical error only the lowest 6 pairs were perfectly matched to not be noticeable with rod-vision.

### Clinical assessments

#### Best corrected visual acuity (BCVA)

BCVA was measured monocularly using ETDRS charts. Eyes were tested at 4m with lens correction if required at 4m with room lights off, windows covered and chart light switched on. The right eye was tested first with ETDRS chart 1 whilst occluding the left eye and then the left eye tested with ETDRS chart 2 whilst occluding the right eye. If the patient could not read at least 20 letters at 4m the test was repeated at 1m. If the testing was conducted at 4m the ETDRS score was the number of letters correctly identified plus 30. If the testing had to be repeated at 1m the score was the number of letters correctly identified at 4m plus those identified at 1m.

For each participant, the untreated eye served as the reference for the expected test–retest variability in Best-Corrected Visual Acuity (BCVA, ETDRS letters). Changes in each BCVA post-treatment measure was calculated as the difference from the mean of all untreated eye pre-treatment measures. The untreated-eye difference range was then pooled across all participants and all post-treatment time points.

#### Optical coherence tomography

SD-OCT scans were conducted on the Spectralis device (Heidelberg Engineering, Heidelberg, Germany). Three separate baseline scans were conducted to establish a mean baseline prior to ATIMP administration. High resolution volumetric (20° x 20°) SD-OCT scans were analysed for structural retinal changes using the HEYEX2 software by a single grader, MK, twice and then quantitative data averaged. Images were automatically segmented for the different retinal layers and then corrected manually if required. Foveal outer nuclear layer thickness was measured by setting the display ratio to 1:1µm on the transfoveal line scan for more accurate measurement. The measurement was taken at the fovea from the border of the inner limiting membrane and the external limiting membrane. For scans showing foveal hypoplasia (persistence of the inner retinal layers over the fovea), the measurement was taken from the outer plexiform-outer nuclear layer border to the external limiting membrane. Patients with foveal hypoplasia were noted. Transfoveal line scans were also used to grade the integrity of the ellipsoid zone (EZ) according to the published grading system described by (Sundaram et al., 2014. The grades are as follows: 1) a continuous EZ, 2) EZ disruption, 3) EZ absence, 4) presence of hyporeflective zone (HRZ) and 5) outer retinal atrophy with RPE loss.

### Spectral sensitivity

Spectral sensitivity measurements were conducted using a custom Maxwellian-view optical system. Radiance was controlled by a combination of fixed neutral-density filters and a computer-controlled variable neutral-density filter (Rolyn Optics, Covina, CA). Sinusoidal flicker was produced by pulse-width modulation of fast, liquid crystal light shutters (Displaytech, Carlsbad, CA) at a carrier frequency of 400 Hz. A monochromator with a 4 nm half-maximum bandwidth was used to select stimulus wavelengths: 450, 500, 550, 600, and 650 nm (H-10 Jobin Yvon, Longjumeau, France). Observers maintained head position with a dental wax impression (bite bar). A 1 Hz flickering 5.9° diameter target was presented at an eccentricity of 5° in the superior retina against a dark background. Full details of the apparatus have been previously published (Stockman et al., 2007; Stockman & Sharpe, 2000).

Measurements were made during the cone plateau, a period following an intense bleach when only cones mediate vision. The bleach consisted of a 30-s exposure to a white Ganzfeld (full field) of 5.50 log_10_ scotopic trolands, which bleaches over 60% of rod photopigment and typically induces a cone plateau lasting between 3-10 minutes post bleach. The cone plateau is characterized by a stable sensitivity with the spectral properties of cone vision. For each trial, participants adjusted the radiance of a continuously flickering target until it was just visible, indicating the threshold. Perceptual judgments were made in runs, with each run comprising three consecutive judgments for each wavelength. A keypad was provided for fine (±0.02 log₁₀ unit) and coarse (±0.10 log₁₀ unit) adjustments.

Participant ACH4 completed 5 runs with their treated eye and 1 run with their untreated eye.

### Critical flicker frequency

Critical flicker frequency measurements were conducted using a custom Maxwellian-view optical system. In brief, the experimental conditions were chosen to favour measurements of the temporal properties of the long-wavelength sensitive (L-) or middle-wavelength sensitive (M-) cones. A flickering target of 4° of visual angle in diameter and either 650 or 588/9 nm in wavelength was presented in the centre of a 9° diameter background field of 480 nm. Fixation was central. The 480-nm background, which delivered 8.26 log quanta s ^−1^ deg ^−2^ at the cornea, served primarily to suppress the rods. In addition, the wavelength of the primary target (650 nm) was chosen to favor detection by cones rather than rods. The secondary target wavelength of 588/9 nm was used to determine the relative spectral sensitivity of the mechanism mediating flicker detection at 650 and 588/9 nm. We use the intensity units of quanta per second per degree squared (quanta s^−1^ deg^−2^). Sinusoidal modulation was produced by the pulse-width modulation of fast, liquid crystal light shutters (Displaytech) at a carrier frequency of 400 Hz. Observers maintained head position with a dental wax impression (bite bar). These systems are described in more detail elsewhere (Stockman et al., 2021). Participants made perceptual judgements on target stimuli at different mean levels of radiance intensities. Approximately 0.3 log unit steps from 6 to 10.3 for 650 nm, and 5.3 to 9.6 log quanta per second per degree squared for 588 nm.

Measurements were made to determine the critical flicker frequency (CFF) threshold. For each trial, participants were presented with a target flickering at 92% contrast (the maximum possible in this optical system) at a given radiance and adjusted its rate of flicker until the flicker was just visible, indicating the threshold. Perceptual judgments were made in runs covering the full range of radiances with a single spectral target (650nm or 588/9nm), with each run comprising three consecutive judgments for each radiance level. Participants were allowed unlimited time to identify this threshold.

ACHM4 completed 3 runs with his treated eye and 3 runs with his untreated eye for each target (588nm and 650nm).

### Rayleigh matches

Stimuli were presented as a 2° bipartite circular field using an HMC Anomaloscope (Oculus, Wetzlar, Germany). The bottom semi-circle (reference field) consisted of a monochromatic 589 nm yellow light. The top semi-circle (test field) comprised an additive mixture of 666 nm (red) and 549 nm (green) primary lights.

For observers with normal trichromatic vision, a perceptual match is only possible when the red-green mixture ratio and the yellow luminance intersect at a point at which the mixture and yellow fields produce the same L- and M-cone excitations (Fig. 5B). For an observer relying on achromatic (rod-based) vision, matches can be found across the full range of red-green ratios whenever the red-green mixture and yellow fields produce the same rod excitations (the rod range; Fig. 5B).

During the experiment, the yellow reference luminance was pre-set to one of four fixed levels. Participants were instructed to adjust only the red-green mixture ratio until the top field perceptually matched the bottom. If typical L/M colour vision has recovered, a match should only be achievable at a single luminance level; matches across multiple luminance levels would indicate a continued reliance on an achromatic mechanism.

## Data Availability

All data produced in the present study are available upon reasonable request to the authors

## Acknowledgments

We thank Nadine Dijkstra, Matan Mazor and Marcus Daghlian for providing comments on an earlier version of this manuscript.

## Author contribution

**R.O.M.M:** Conceptualization, Methodology, Analysis, Investigation, Writing - Original Draft;

**M.F**: Methodology, Analysis, Investigation;

**A.T.R:** Methodology, Analysis, Investigation, Writing - Review/Editing

**A.S:** Methodology, Writing - Review/Editing;

**T.B.P:** Investigation, Writing - Review/Editing;

**M.M.K**: Investigation;

**M.M**: Conceptualization, Methodology, Writing - Review/Editing

**T.M.D**: Conceptualization, Methodology, Investigation, Writing - Original Draft;

## Supplementary Materials

Supplementary materials are available at the end of this document, located after the references.

## Supplement 1 Results of the cone-selective fMRI stimuli psychophysics

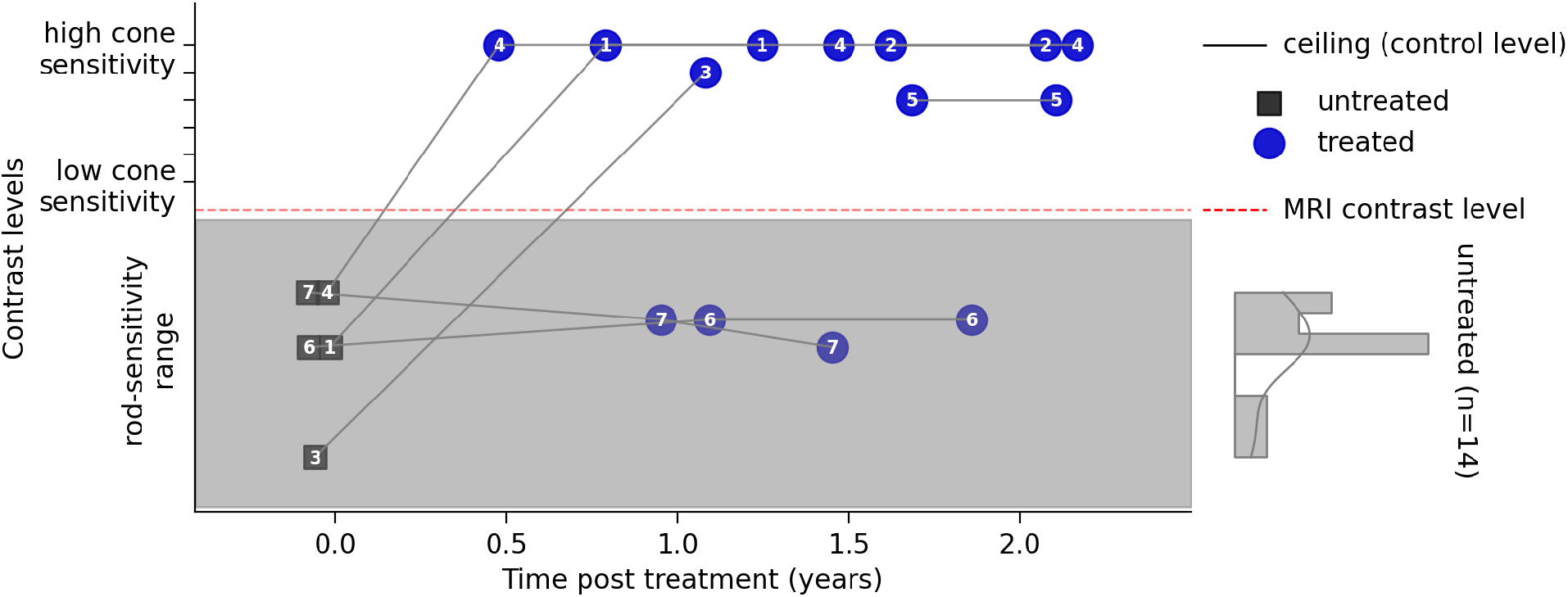

To objectively test the conscious perception of the stimulus in the scanner, we measured binocular cone contrast sensitivity thresholds for the cone-selective ring-and-wedge stimuli, identical to that used during the cone-selective pRF mapping runs. In a 2AFC task (2 Alternative Force Choice), participants detected movement of a checkerboard ring (inward/outward) or wedge (clockwise/anticlockwise) reversing at 2 Hz. Both an incorrect response and a report that no checkerboard was visible after multiple replays triggered a step up in cone contrast, while a correct response triggered a step down in cone contrast (1-up/1-down adaptive staircase). Each 2AFC 1-up/1-down adaptive staircase continued until at least 8 reversals (mean N trials = 25), or >8 correct responses had occurred at the dimmest contrast level (ceiling). For each eye and patient, the resulting threshold (on the Y-axis) is one of twenty-one contrast levels, generated using silent-subtitution as equidistant steps between the highest and lowest steps achievable in our setup. Unfortunately due to a technical error only the lowest 6 pairs were perfectly matched to not be noticeable with rod-vision.

## Supplement 2 Polar angle maps of cone-selective stimuli

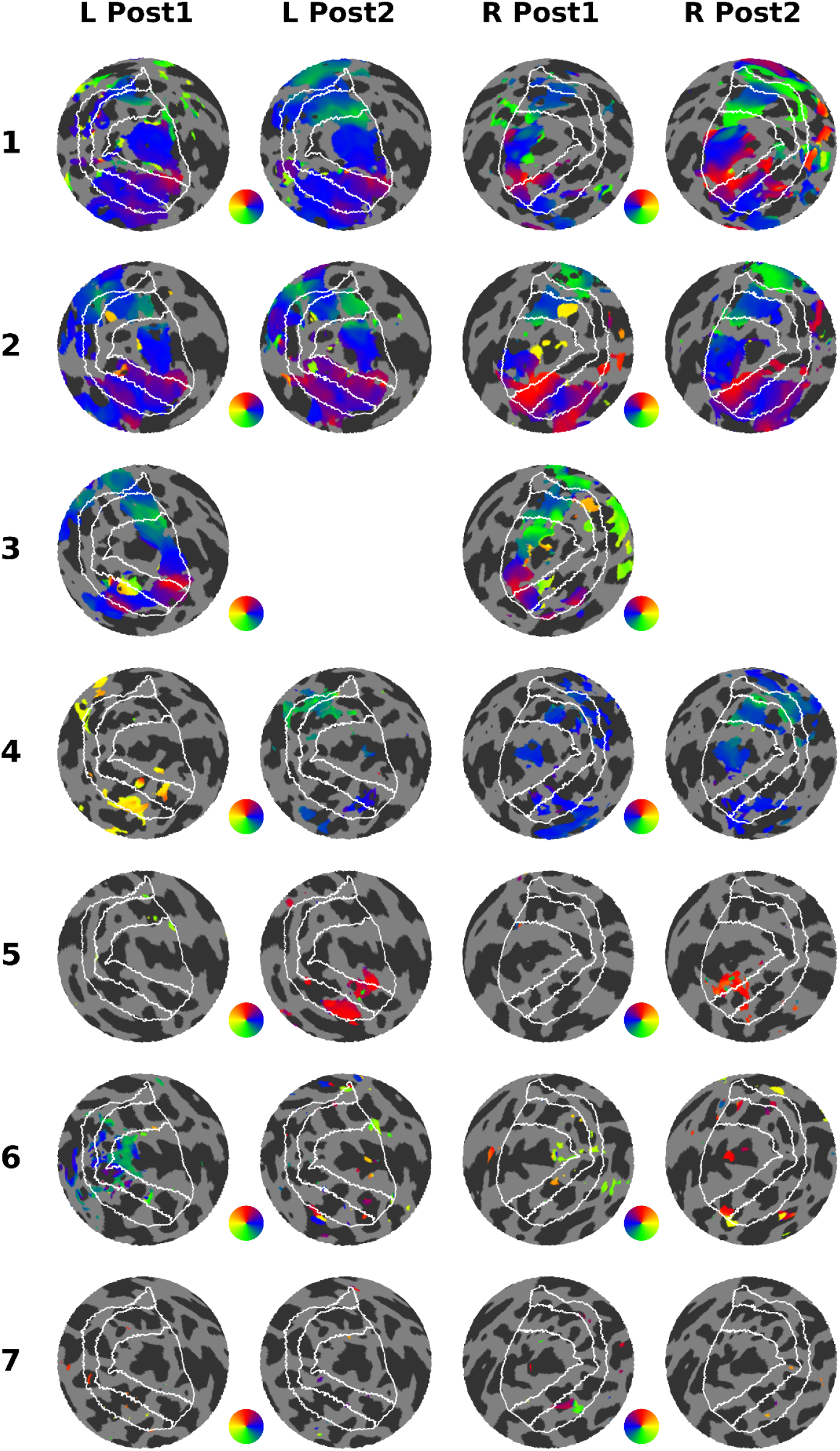

## Supplement 3 Detailed report of fMRI measures per session

**Table S3.** Coverage of cone-selective signals in V1-3 in percentage of vertices in the ROI with R2>0.1, across sessions

| patient | pre | post1 | post2 |
| --- | --- | --- | --- |
| ACH1 | 3.5 | 42.7 | 61.0 |
| ACH2 | 0.4 | 49.6 | 56.2 |
| ACH3 | 0.7 | 41.5 | NaN |
| ACH4 | 0.0 | 11.8 | 19.9 |
| ACH5 | 13.5 | 0.5 | 7.6 |
| ACH6 | 1.7 | 10.0 | 3.1 |
| ACH7 | 7.8 | 1.5 | 0.4 |

## Supplement 4 Rod retinotopy maps

Eccentricity and polar angle maps of pRF mapping for left and right hemispheres using rod-selective stimuli in the two post treatment sessions. Showing that the lack of recovery in ACH5-7 was not due to insufficient data quality in these patients.

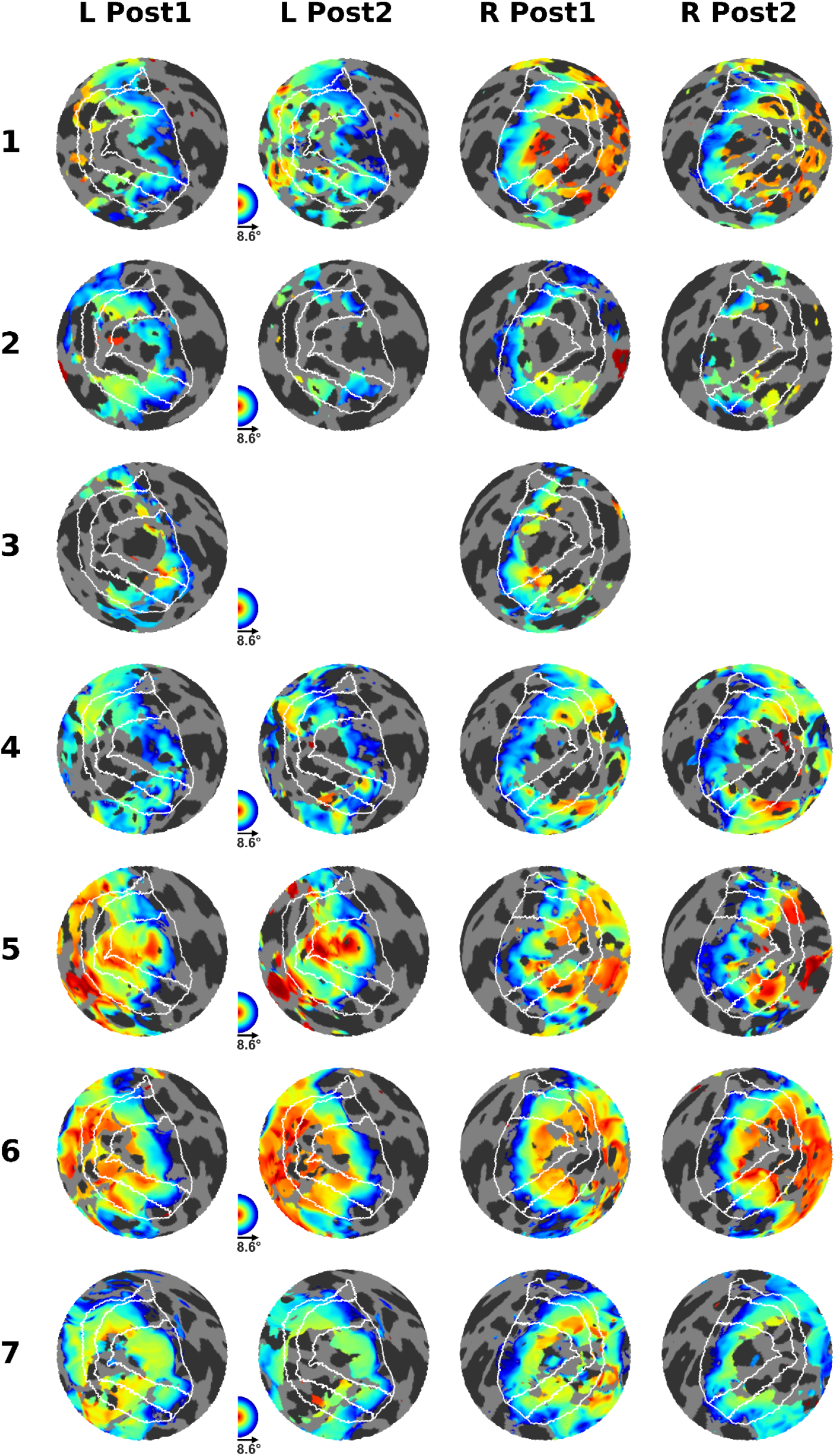
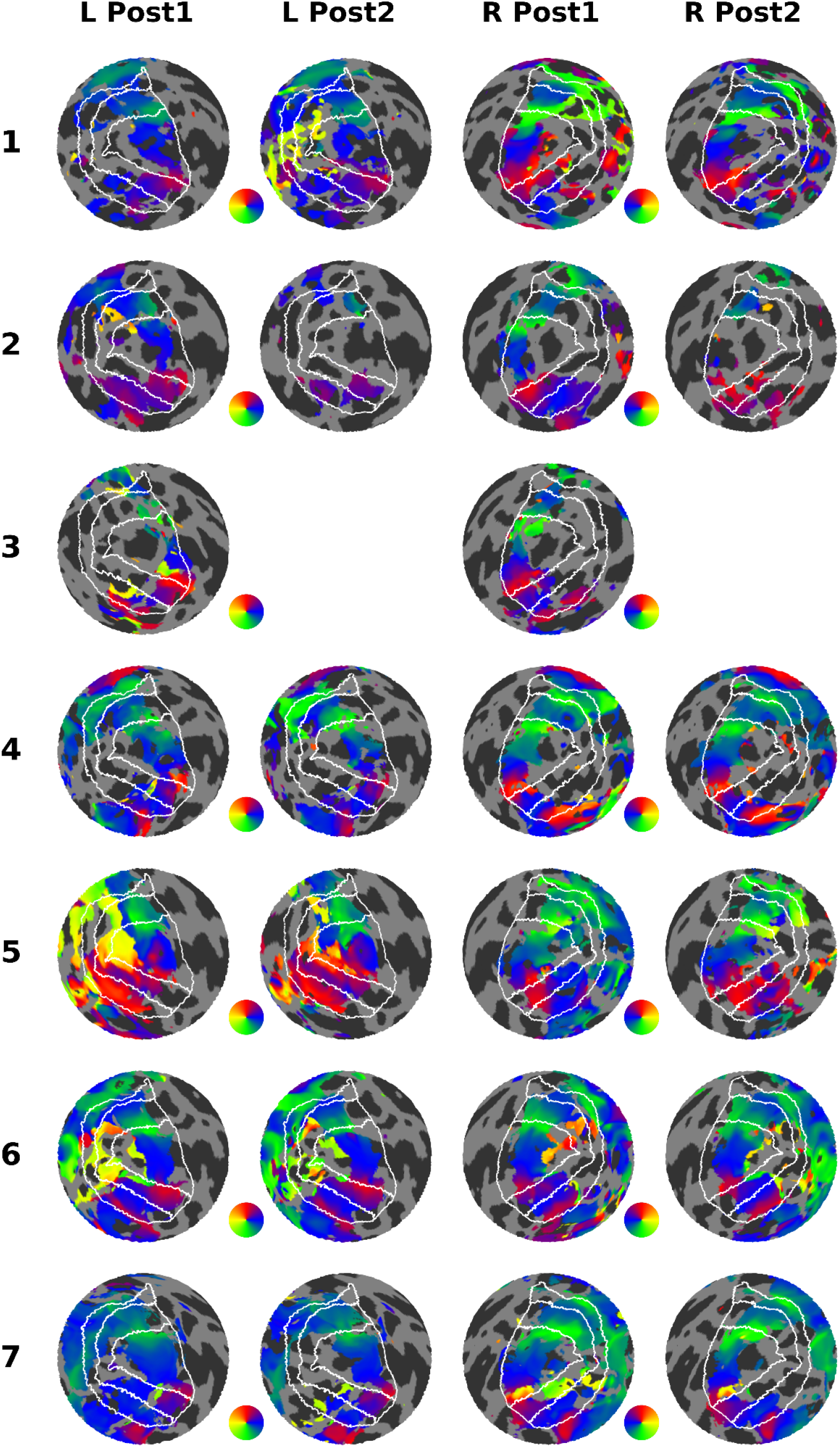

## Supplement 5 Additional Colour tests

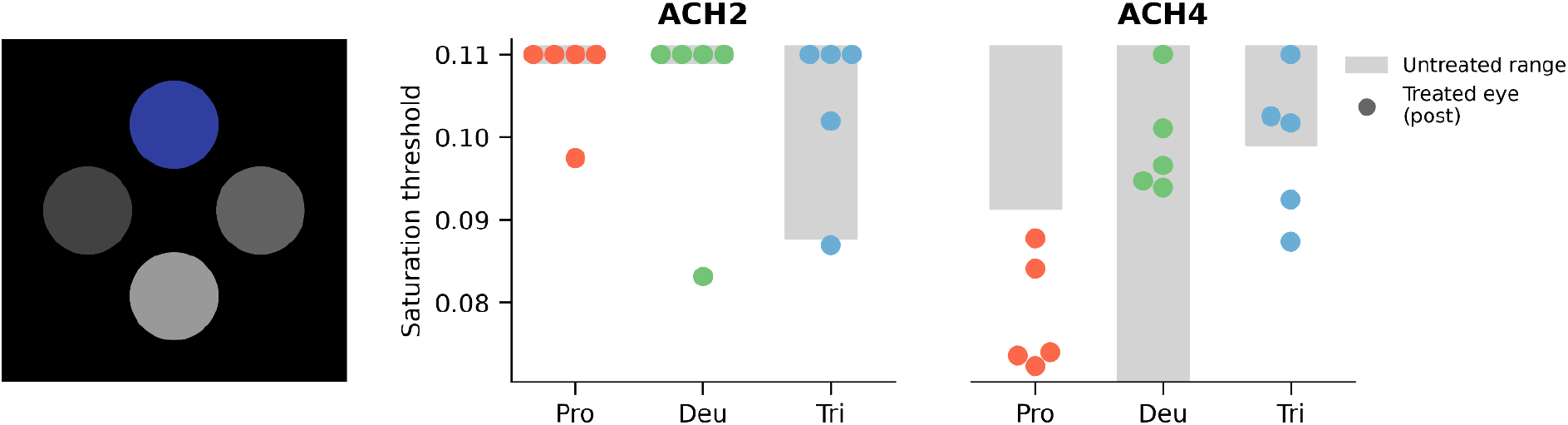

### Low-vision version Cambridge colour test (Metropsis)

In the low-vision version Cambridge colour test, participants are asked to pick out which of four circles is coloured (chromatic) and thus distinguish it from three monochrome “grey” (achromatic/neutral hue) circles. The task has three variations that test for colour discrimination along the protan, deutan and tritan confusion lines that diagnose anomalies in the L-, M- and S-cones respectively. A saturation threshold of 0.11 means no colour perception. A person with normal trichromat vision will have a threshold of ∼0.01.

Depending on which cones have recovered and which post-photoreceptor mechanisms have recovered we might expect recovery in all, some or none of these conditions. ACH2 showed no consistent improvement while ACH4 showed a consistent improvement along the protan axis.

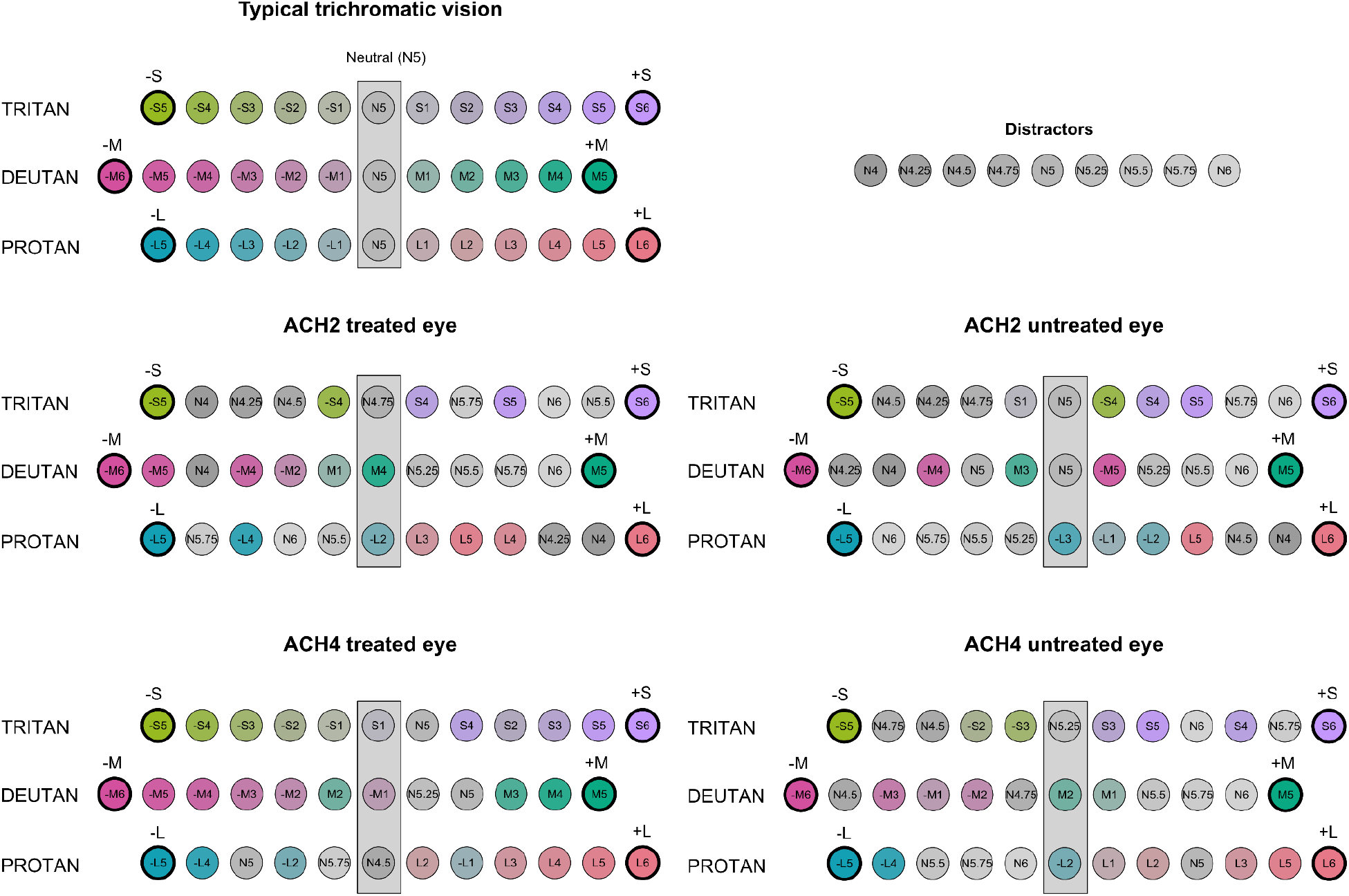

### Stockman Colour Test

In the Stockman colour test, participants were presented separately with three sets of coloured and monochromatic discs aligned along tritan (-S5 to S6), deutan (-M6 to M5)and protan (-L5 to L6) confusion lines that passes through a grey (N5) to assess S-, M- and L-cone deficits, respectively. The discs varied steadily in colour between the two extreme hues (-S5 and S6,-M5 and M5, and -L5 and L6). The experimenter mixed up the discs, including the grey distractors (N4 to N6) and asked participants to arrange them in colour order between the two extremes. In line with the Rayleigh matching results, ACH2 showed poor chromatic discrimination in his treated and untreated eye and frequently included achromatic distractors, but perhaps showed marginally better discrimination in the +S, -M and +L directions. ACH4 showed a similarly poor performance in his untreated eye, but performed much better in his treated eye in all three directions, making fewer ordering and performing much closer to a normal control. His performance in this test is the first evidence that suggests some recovery in S-cone performance along the tritan direction. Interestingly, while he reports that L-cone increments and M-cone decrements appear more “red/warm” similar to a normal control, he also reports S-cone decrements appear “more red/warm” compared to increments.

